# A consensus Covid-19 immune signature combines immuno-protection with discrete sepsis-like traits associated with poor prognosis

**DOI:** 10.1101/2020.06.08.20125112

**Authors:** Adam G. Laing, Anna Lorenc, Irene Del Molino Del Barrio, Abhishek Das, Matthew Fish, Leticia Monin, Miguel Muñoz-Ruiz, Duncan R. McKenzie, Thomas S. Hayday, Isaac Francos-Quijorna, Shraddha Kamdar, Magdalene Joseph, Daniel Davies, Richard Davis, Aislinn Jennings, Iva Zlatareva, Pierre Vantourout, Yin Wu, Vasiliki Sofra, Florencia Cano, Maria Greco, Efstathios Theodoridis, Joshua Freedman, Sarah Gee, Julie Nuo En Chan, Sarah Ryan, Eva Bugallo-Blanco, Pärt Peterson, Kai Kisand, Liis Haljasmägi, Lauren Martinez, Blair Merrick, Karen Bisnauthsing, Kate Brooks, Mohammad Ibrahim, Jeremy Mason, Federico Lopez Gomez, Kola Babalola, Sultan Abdul- Jawad, John Cason, Christine Mant, Katie J Doores, Jeffrey Seow, Carl Graham, Francesca Di Rosa, Jonathan Edgeworth, Manu Shankar-Hari, Adrian C. Hayday

**Affiliations:** Peter Gorer Department of Immunobiology, School of Immunology and Microbial Sciences, King’s College London, London, UK; The Francis Crick Institute, London, UK; UCL Cancer Institute, University College London, London, UK; London School of Hygiene & Tropical Medicine, London, UK; Regeneration Group, Wolfson Centre for Age-Related Diseases, IoPPN, King’s College London, London, UK; Department of Plastic and Reconstructive Surgery, Royal Free NHS Foundation Trust, London, UK; Department of Inflammation Biology, King’s College London, London, UK; Molecular Pathology Research Group, Institute of Biomedicine and Translational Medicine, University of Tartu, Tartu, Estonia; Infectious Diseases Department, Guy’s and St Thomas’ NHS Foundation Trust, London, UK; Immunological Medicine, King’s College Hospital NHS Foundation Trust, London, UK; The European Bioinformatics Institute (EMBL-EBI) Wellcome Genome Campus, Hinxton, UK; Comprehensive Cancer Centre, School of Cancer & Pharmaceutical Sciences, King’s College London, London, UK; Department of Infectious Diseases, School of Immunology and Microbial Sciences, King’s College London, London, UK; Infectious Diseases Biobank, Department of Infectious Diseases, School of Immunology and Microbial Sciences, King’s College London, London, UK; Institute of Molecular Biology and Pathology, National Research Council of Italy (CNR), Rome, Italy; Centre for Clinical Infection and Diagnostics Research, Department of Infectious Diseases, Guy’s and St Thomas’ NHS Foundation Trust, London, UK; Department of Intensive Care Medicine, Guy’s and St Thomas’ NHS Foundation Trust, London, UK

## Abstract

Person-to-person transmission of SARS-CoV-2 virus has triggered a global emergency because of its potential to cause life-threatening Covid-19 disease. By comparison to paucisymptomatic virus clearance by most individuals, Covid-19 has been proposed to reflect insufficient and/or pathologically exaggerated immune responses. Here we identify a consensus peripheral blood immune signature across 63 hospital-treated Covid-19 patients who were otherwise highly heterogeneous. The core signature conspicuously blended adaptive B cell responses typical of virus infection or vaccination with discrete traits hitherto associated with sepsis, including monocyte and dendritic cell dampening, and hyperactivation and depletion of discrete T cell subsets. This blending of immuno-protective and immuno-pathogenic potentials was exemplified by near-universal CXCL10/IP10 upregulation, as occurred in SARS1 and MERS. Moreover, specific parameters including CXCL10/IP10 over-expression, T cell proliferation, and basophil and plasmacytoid dendritic cell depletion correlated, often prognostically, with Covid-19 progression, collectively composing a resource to inform SARS-CoV-2 pathobiology and risk-based patient stratification.

---

The SARS-CoV-2 coronavirus is the aetiologic agent of Covid-19, a catastrophically disruptive pandemic^1,2^. Most infected individuals recover, with many remaining pauci- or asymptomatic. Nonetheless, vast numbers of infected persons have required hospitalisation for severe respiratory distress together with variable complications including a chronic hyperinflammatory state^3,4^, and hundreds of thousands have died^5^.

In general, limiting virus-triggered disease and death is achieved by developing antivirals, vaccines, and/or immune-modulators, for which empirical and/or repurposing approaches can be successful. Nonetheless, such strategies are best informed by understanding the host-pathogen relationship that in the case of SARS-CoV-2 is likely to blend unique facets with those common to other coronaviruses^6^. Hence, understanding the host-pathogen relationship may inform ongoing clinical care, and increase preparedness for potential SARS-CoV-2 resurgence, and/or for the possible emergence of other highly pathogenic coronaviruses^7^.

The immune response is a core component of host-pathogen relationships. Its understanding can identify correlates of protection, that can vary greatly in different settings, and correlates of pathology, sometimes reflecting disease-promoting immune dysregulation mechanisms that are common to diverse clinical manifestations. For SARS-CoV-2, the full recovery of most infected individuals probably reflects their mounting protective B and T cell responses^8,9^. Likewise, several patients presenting with SARS disease in 2002-2003 showed SARS-CoV-1-specific antibodies and T cells some years later^10,11^. Additionally, vaccines have been developed to prevent severe coronavirus-induced diseases of pigs, chickens, and dogs, albeit with variable efficacy^12^.

In this context, infected persons requiring hospitalization for Covid-19 disease may reflect deficiencies in generating virus-neutralising antibodies, in which regard patients have been transfused with antibody-rich plasma from convalescent subjects^13,14^. Moreover, given that the elderly make poor antigen-specific responses to seasonal influenza vaccination^15^, relative immunodeficiency might partly explain age-related increases in Covid-19. This notwithstanding, hospitalization might reflect exaggerated immune responses compounding virus-induced damage to promote vasculopathy and organ dysfunction *via* “cytokine storms”. Thus, SARS1 patients were administered immuno-suppressants^16^, and a cytokine-neutralising antibody, anti-IL6, has been deployed in Covid-19^6,17,18^. It is likewise plausible that different Covid-19 manifestations reflect different positions on a spectrum of immunodeficiency and immunopathology, and that single individuals may traverse this spectrum as their disease progresses through discrete phases^19^. Additionally, contributions to disease susceptibility of age, gender, and genetics, and of conditions such as hypertension, diabetes, and respiratory illness^20-22^ may partly reflect their impacts on immuno-protective and/or immunopathological mechanisms.

Immunological studies of SARS-CoV-2 infected persons, particularly those hospitalised, have reported immunological responsiveness, and apparent disease-associated immune deficits, including sub-optimal Type-I and Type-III IFN responses, excessive activation of lung-infiltrating neutrophils, pan-lymphopenia, myeloid cell dysregulation, and T and NK cell exhaustion^6,23-27^. Combining these important findings with precedents set by SARS1, MERS, and animal coronavirus infections, we hypothesised that despite their heterogeneity, persons hospitalised for Covid-19 might display a consensus core immune signature, just as highly diverse individuals display consensus vaccine response signatures^28^.

In seeking this signature, we have used two frames-of-reference, each informed by high-throughput, multicomponent analyses of human immune responses: first, immuno-protective responses induced by acute virus infection or vaccination^28-30^; second, sepsis as an established example of potentially life-threatening illness fuelled by dysregulated immunoprotective responses, including neutrophil hyperactivation, atypical monocyte activation, reduced natural killer (NK) and dendritic cell (DC) function, and concomitant activation, suppression and depletion of B and T lymphocytes^31^. Although the clinical presentation of Covid-19 differs from sepsis, we hypothesised that a Covid-associated immune signature may share distinct traits with the sepsis immunophenotype. Identifying those traits might reveal key aspects of the SARS-Cov-2-host relationship, potentially guiding much-needed treatment strategies.

To test this hypothesis, we sampled peripheral bloods of 63 Covid-19 patients treated at Guy’s and St Thomas’ Trust Hospitals and of 55 co-analysed healthy controls (HC), among whom a small minority were exposed-but-recovered, non-hospitalised subjects. The study is termed Covid-IP (Covid-ImmunoPhenotyping), and the hospitalised Covid-IP patient cohort is termed CP. Although Covid-19 is mostly defined by respiratory failure contributed to by local inflammation, and frequently combined with coagulopathies and other organ failures, peripheral blood sampling has many merits, specifically: its practicality in multiple settings and locations thereby facilitating meta-analyses embracing scenarios such as vaccination and sepsis, and comparison with routine clinical blood measurements^29,31^; and its established capacity to reflect, albeit incompletely, immune cells trafficking to and from tissues, particularly when appropriate experimental approaches are employed to maximise that capacity^32,33^.

The data obtained strongly support the hypothesis that across a highly variable cohort, a consensus Covid-19 immune signature was discernible in peripheral blood. The signature captured several aspects of previously reported Covid-19 immunophenotypes, blending specific traits of vaccine responses and sepsis, respectively, with less commonly cited traits, including major depletions of blood plasmacytoid dendritic cells (pDC) and basophils, and overtly high cycling of effector-memory CD4^+^ and CD8^+^ T cells concomitant with selective T cytopenia. Moreover, some traits correlated, often prognostically, with evolving disease severity. Among those was sustained CXCL10/IP10 (hereafter “IP10”) overexpression as occurs in SARS1 and MERS. In short, the core Covid-19 immune signature provides a resource offering specific insights into the SARS-CoV-2-host relationship, and practical prospects of a prognostic disease signature to aid risk-based patient stratification. If the immunological signature not only tracks but contributes Covid-19, our findings support therapeutic strategies to boost T cell competence, e.g. by use of IL7, currently on trial at St Thomas’ Hospital, possibly combined with IP10 antagonism.

## Results

### Covid-IP

Many methods facilitate human immune monitoring, including mass cytometry and single cell RNA-sequencing^34,35^. In designing Covid-IP we sought high-throughput standard-operating-protocols that could be easily applied globally, thereby increasing opportunities for Covid-19 meta-analysis. Thus, we applied eight multiparameter flow cytometry panels (P1-P8) that measured: [1] broad lymphocyte composition; [2] effector/memory T cell status; [3] γδ T cell status; [4] B cells; [5] cell cycling; [6] absolute leukocyte counts; [7] lymphocyte activation and exhaustion; and [8] innate immune cells. P1-P5 were applied to peripheral blood mononuclear cells, and P6-P8 to whole blood. Gating strategies are described and illustrated in **Supplementary Materials**. Also measured were 22 cytokines, and antibodies against SARS-CoV-2 nuclear capsid (N), spike (S), and receptor binding domain (RBD). Thereby, ∼600 data points were obtained from each sample.

The CP cohort was compared with the comparably-sized HC cohort (**Supp Table 1**) by values of individual parameters; by the distribution of values, as reflected in coefficients of variation (CV); and by the overall immunological structures of CP and HC which reflect aggregate correlations between discrete parameters. Where relevant, potential impacts of age and gender were corrected for prior to making statistical CP vs HC comparisons (**Supp Table 2**). Because, the timing, anatomical route, dose, multiplicity and nature of infection were unknown, and the declared onset of symptoms unreliable, the only point of certainty to be denoted as Day 1 was within 24h of in-patient identification as SARS-CoV-2^(+)^, which occurred from 1-15 days following declared symptom-onset. All patients were sampled on Day 1; approximately two-thirds on Day 3; and, those remaining hospitalised were sampled again on Day 9.

To classify disease severity, we re-purposed an eight-point ordinal scale recommended by the World Health Organisation (WHO) Research and Development Blueprint Expert group for classifying Covid-19 trial endpoints: “low” (WHO scores 1-2) [6 patients] reflected mild Covid-19 symptoms of patients who scored as SARS-CoV-2^(+)^ but were hospitalised for other reasons; “moderate” (WHO scores 3-4) [26 patients] reflected symptoms but little or no requirement for supplemental oxygen; and “severe” [31 patients] reflected a disease course including any or all of: high-flow oxygen requirement (score 5), mechanical ventilation (6), multi-organ support (7), and death (score 8). Within the severe group, four were hyperinflammatory (HI), manifested by persistent fevers and hypoxaemia, hyperferritinaemia, high CRP, and cytopenia, in combination with negative bacterial cultures. Importantly, scores represent the peak severities for each patient: hence, immunological parameters that on Day1 correlated with severity were at minimum associative and commonly prognostic. Additionally, four patients presented with possibly confounding factors (denoted “CF”; **Supp Table 1**) (e.g. treatment with a monoclonal antibody antagonising B cell activation factor; and a JAK2 kinase gain-of-function mutation) which were noted because of their potential to skew HC *vs* CP comparisons.

Principal component analysis of 147 flow cytometry phenotypes for subjects for whom full data-sets were available very clearly segregated CP from HC (**Fig 1a**). Among individual changes, CP displayed high levels of plasma IP10, IL6, and IL8; CD4^+^ and CD8^+^ effector-memory (T_EM_) cells in the G1 phase of the cell cycle, and anti-RBD IgG, in the context of overt T cytopenia (**Fig 1b**). Significant positive and negative correlations between discrete parameters (that do not reflect reciprocal co-dependencies) collectively impose distinct structures on immune systems in different settings^36^. To compose a Covid-IP immune structure (Fig 1c), the correlations used were those with R values ≥0.3 above or below those that characterise HC. These included exaggerations of HC correlations; novel correlations for which one criterion does not usually exist in HC (e.g. IP10 levels vs CXCR3^+^CCR6^neg^CD8^+^ cells, or anti-RBD IgG *vs* naive CD4^+^ T cells); and inversions of HC relationships (e.g. basophils vs CD4^+^ T cells, or CD4^+^ T effector memory (T_EM_) cells vs PD1^+^ Vδ1^+^ T cells) (**Fig 1d, e**).

**Figure 1:**
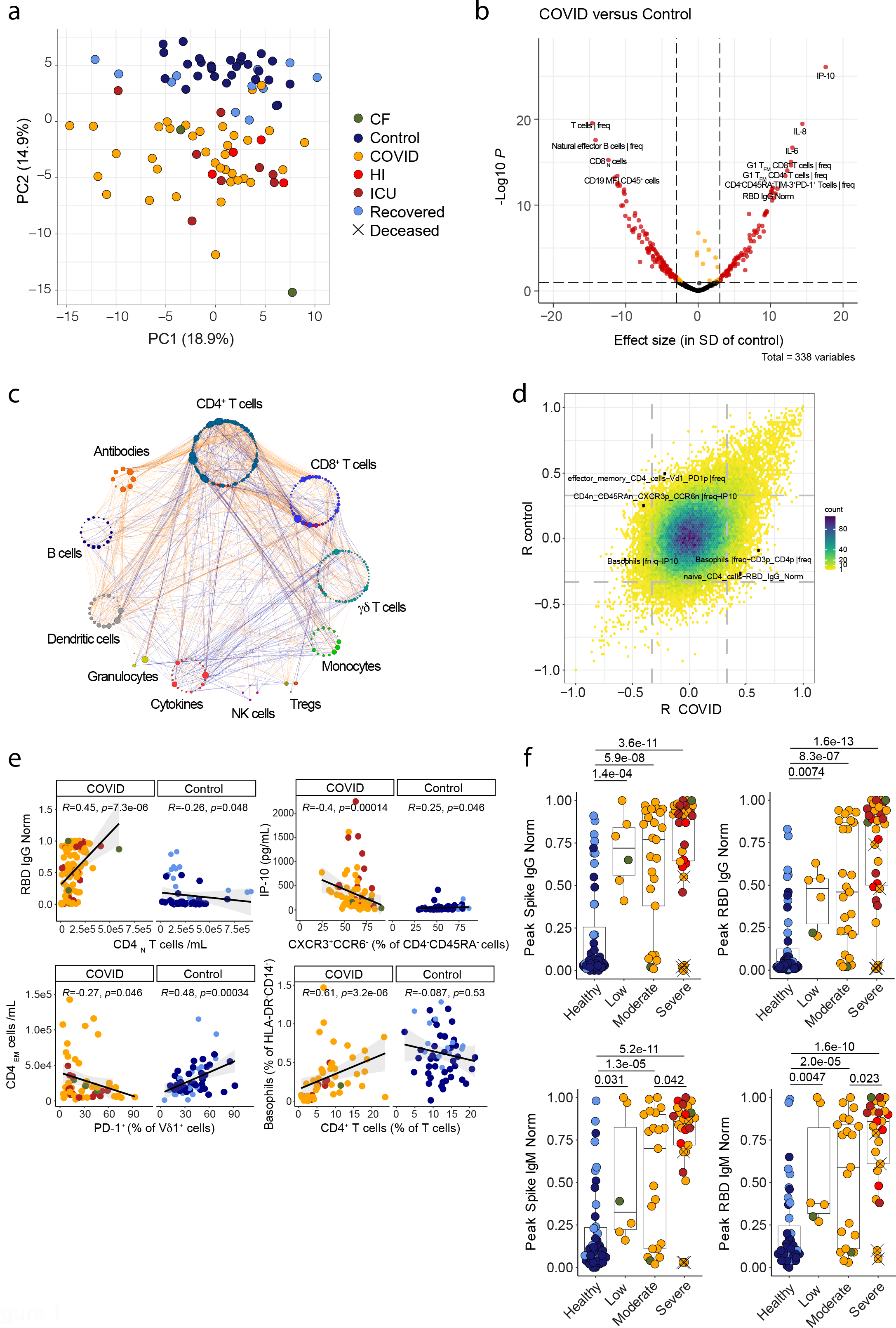
Immunophenotyping reveals distinct features of the immune system in COVID-19 patients. **a)** Principal component analysis of 147 cell type frequencies analysed in COVID and control donors. PC1 explains 18.9% of the variation while PC2 explains 14.9% of the variation; colour denotes disease status. n=92 n’=74. **b)** Volcano plot of 338 non-redundant immune parameters analysed in COVID relative to control samples. Parameters significantly affected in COVID patients (p<0.01, difference of means at least 3 SD relative to the control group) are shown in red. Control n=78 n’=55, COVID n=114 n’=63 **c)** Spearman correlation network in COVID samples (n=114, n’=63). Nodes are manually clustered into functional groups of immune parameters. Node size corresponds to the degree of relatedness. Edge colour denotes direction of correlation, orange = positive correlation, navy = negative correlation. Only Spearman correlations in which p<0.01, R≥0.3 or < -0.3 and a delta R of greater than +/-0.3 relative to Control (n=78,n=55) values are represented. **d)** Correlations between immune parameters in control (y axis) and COVID patients (x axis). Colour corresponds to the number of overlapping points. Note that all points outside the dashed vertical and horizontal lines at R -0.33 and 0.33 denote significant correlations (p<0.01). **e)** Correlations between a subset of parameters with significantly different correlation coefficients in COVID relative to control. Plots display results from Spearman correlation tests and a linear regression line with 95% confidence interval shading. **d-e)** Control n=78 n’=55, COVID n=114 n’=63. **f)** Peak antibody titres against SARS-CoV-2 Spike and RBD antigens. Healthy n’=43, Low n’=6, Moderate n’=21, Severe n’=25. n=samples, n’=individuals. n/n’ may vary slightly between graphs due to data filtering or experimental dropouts (see methods). Box plots denote median and 25^th^ to 75^th^ percentiles (boxes) and 10^th^ to 90^th^ percentiles (whiskers) and were statistically evaluated by a linear mixed model grouped by severity, with patient as random variable, corrected for age- and sex-dependency. CF: confounding factors; HI: hyperinflammatory; ICU: intensive care unit.

Almost all CP patients displayed high levels of circulating anti-SARS-CoV-2 IgM and IgG antibodies, as did several HC subjects exposed to SARS-CoV-2 prior to study commencement, and not requiring hospitalisation (**Fig 1f; Supp Fig 1a,b**). Antibodies were detected independently by ELISA and by Luciferase-based Immunoprecipitation (LIPS) assays that correlated well (**Supp Fig 1c**). Of note, three of five patients who died during Covid-IP failed to make anti-SARS-CoV-2 IgG, but in two cases death occurred within 3-5 days of symptom- onset, prior to the time-window probably required for antibody production.

### Robust B cell responses to SARS-CoV-2

Given the near-universal development of antibodies in CP, we next examined their B cells. Median values of overall B cell numbers were significantly albeit slightly reduced in CP vs HC, but inter-individual variation within CP was such that the cohort embraced small groups with overt cytopenia (<10^4^ B cells/ml) and atypically high values (2-3×10^5^/ml), respectively (**Fig 2a**). Interestingly, B cells across CP showed a consensus reduction in CD19 expression levels (**Fig 2b**), together with signature shifts in subset composition; e.g. a significant reduction in CD5^+^ B cells, that ordinarily account for ∼25% of B cells, but which in CP often reflected <10% (**Fig 2c**). Similarly, overt reductions occurred in CD27^+^IgM^+^IgD^+^ cells, commonly regarded as “natural effector” cells (**Supp Fig 1d**). Unsurprisingly, CP showed a significant although slight reduction in naïve IgM^+^CD27^neg^ B cells (**Fig 2d**), *versus* significantly increased CD38^+^CD27^+^ plasmablast counts, although as for total B cells, the range was considerable (**Fig 2e**). In part this reflected a transience of human plasmablast expansion, that reportedly peaks 1-2 weeks’ post-vaccination and which was apparent in longitudinal CP samples (**Fig 2f-h**). Likewise, plasmablast expansions were not evident in recovered SARS-CoV-2 seropositive HC individuals (above) (**Fig 2e,g**). In only two cases were (mildly) increased plasmablast frequencies not associated with seroconversion (**Fig 2g**), but possibly because peak plasmablast expansion may often have preceded CP sampling, plasmablast numbers correlated only weakly with S-specific or RBD-specific IgM or IgG (**Fig 2h**).

**Figure 2:**
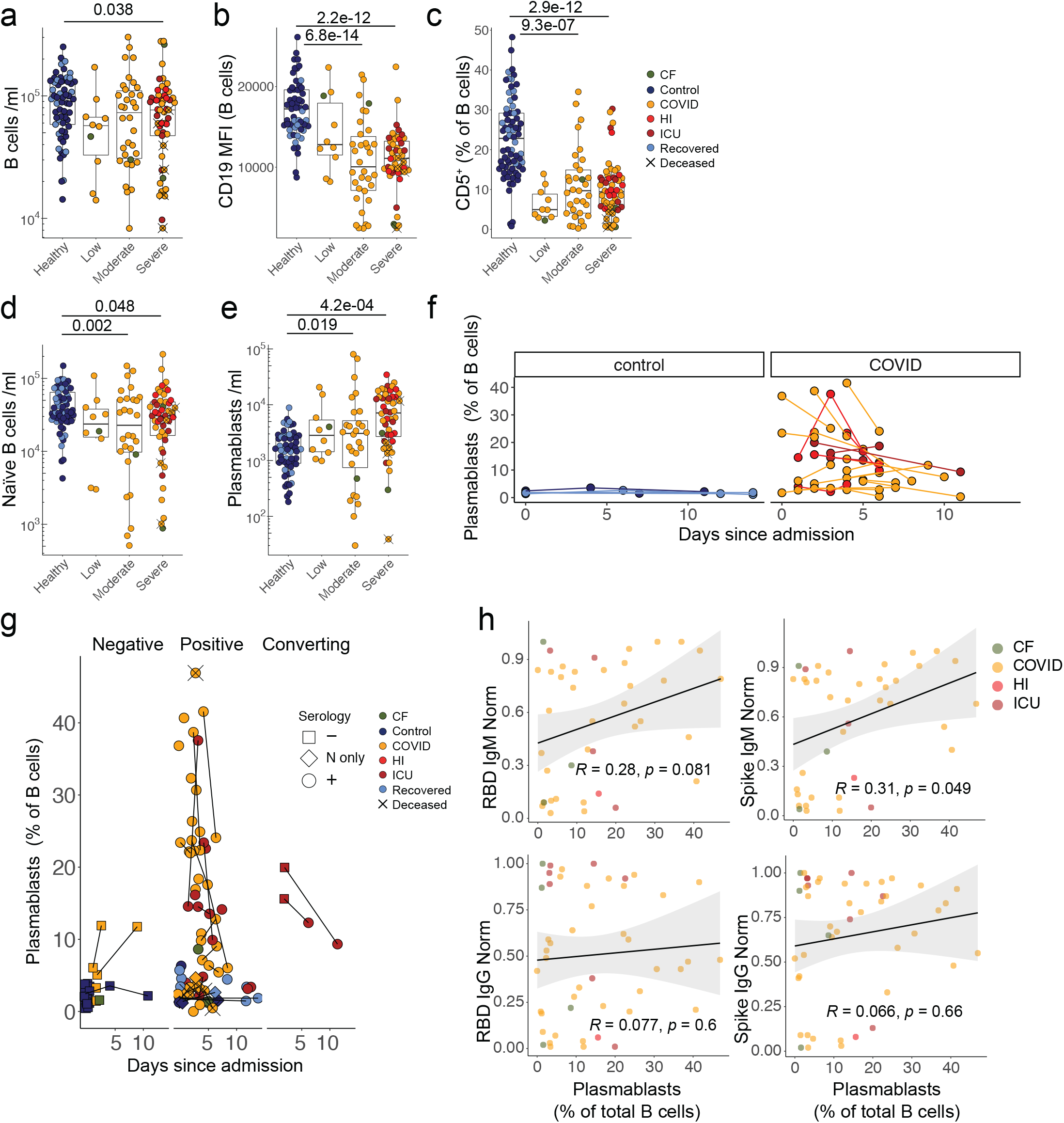
Co-existent anti-SARS-Cov-2 antibody responses and disruption to the B cell compartment in COVID-19 patients. **a)** B cell numbers. **b)** CD19 MFI within B cells. **c)** Frequency of CD5^+^ B cells. Quantification of d) IgM^+^CD27^-^ naïve B cells and **e)** plasmablasts. **a-e)** Healthy n=64 n’=48, Low n=10 n’=6, Moderate n=35 n’=24, Severe n=53 n’=28. **f)** Plasmablast frequencies over time in control and COVID individuals with multiple sampling dates (in samples <15 days after admission). Repeat samples from the same individual are linked. For controls, longitudinal samples are those co-analysed with patients on the denoted days since patient admission. Control n=12 n’=5, COVID n= 50 n’= 23. **g)** Plasmablast frequencies in control and COVID individuals relative to seropositivity for SARS-CoV-2 antigens (N only = IgM positive for N only, + = positive for one or more IgG and/or IgM against N, Spike and/or RBD. Threshold for positive antibody response = 0.3). Repeat samples from the same individual are linked. For controls, longitudinal samples are those co-analysed with patients on the denoted days since patient admission. Negative n=30 n’=26, Positive n=77 n’=57, Converting n=4 n’=2. **h)** Correlation between peak plasmablast frequencies and SARS-CoV-2-specific antibody titres. n = 103, n’ = 62. n=samples, n’=individuals. n/n’ may vary slightly between graphs due to data filtering or experimental dropouts (see methods). Correlation plots display results from Spearman correlation tests and a linear regression line with 95% confidence interval shading. Box plots denote median and 25^th^ to 75^th^ percentiles (boxes) and 10^th^ to 90^th^ percentiles (whiskers) and were statistically evaluated by a linear mixed model grouped by severity (excluding low), with patient as random variable, corrected for age- and sex-dependency.CF: confounding factors; HI: hyperinflammatory; ICU: intensive care unit.

In sum, most CP patients displayed changes in B cell composition resembling those reported following other virus infections or vaccination. Most produced SARS-Cov2-specific antibodies, whose host-protective potentials were inferred from strong correlations of RBD-specific IgG measured by ELISA with a virus-entry neutralisation assay applied to a subset of samples (*K*.*D*., *unpublished*). However, there was no clear correlation of disease severity with either SARS-Cov-2-specific IgM or IgG or with anti-thyroglobulin and thyroid peroxidase antibodies (**Supp Fig 1e**) that were assessed in a small cohort because of their correlations with adverse events following swine-flu vaccination^28^.

### Innate immune cytokines

B cell responses to intramuscular vaccination were preceded by strong innate immune responses detectable in peripheral blood. Those included striking but transient increases in IP10, a chemokine induced by Type-I or Type-II interferon (IFN)^28^. Increased IP10 levels were almost universal across CP, but unlike vaccination, levels were frequently sustained and strikingly proportional to severe disease progression, discriminating severe from moderate and moderate from low (**Fig 3a; Supp Fig 2a**). IP10 levels were correlated with that can induce IP10 expression (**Fig 3b**), but because the CV for IFNγ in HC was high, significant disease-associated increases were largely limited to severe CP (**Supp Fig 2b**). IP10 levels also correlated with IL6 and with IL10 (**Fig 3b**). Nonetheless, while IL6 and IL10 were over-expressed in CP, and correlated with each other, their expression across moderate and severe disease was mostly comparable (**Fig 3c,d**). Likewise, IL8 (CXCL8) was substantially over-expressed across CP, but almost completely unrelated to severity (**Fig 3d**). Beyond those mentioned, other cytokines and chemokines measured failed to significantly discriminate CP from HC (**Supp Fig 2c**). Although a small CP sub-cohort sustained high levels of several cytokines, this was not a consensus signature.

**Figure 3:**
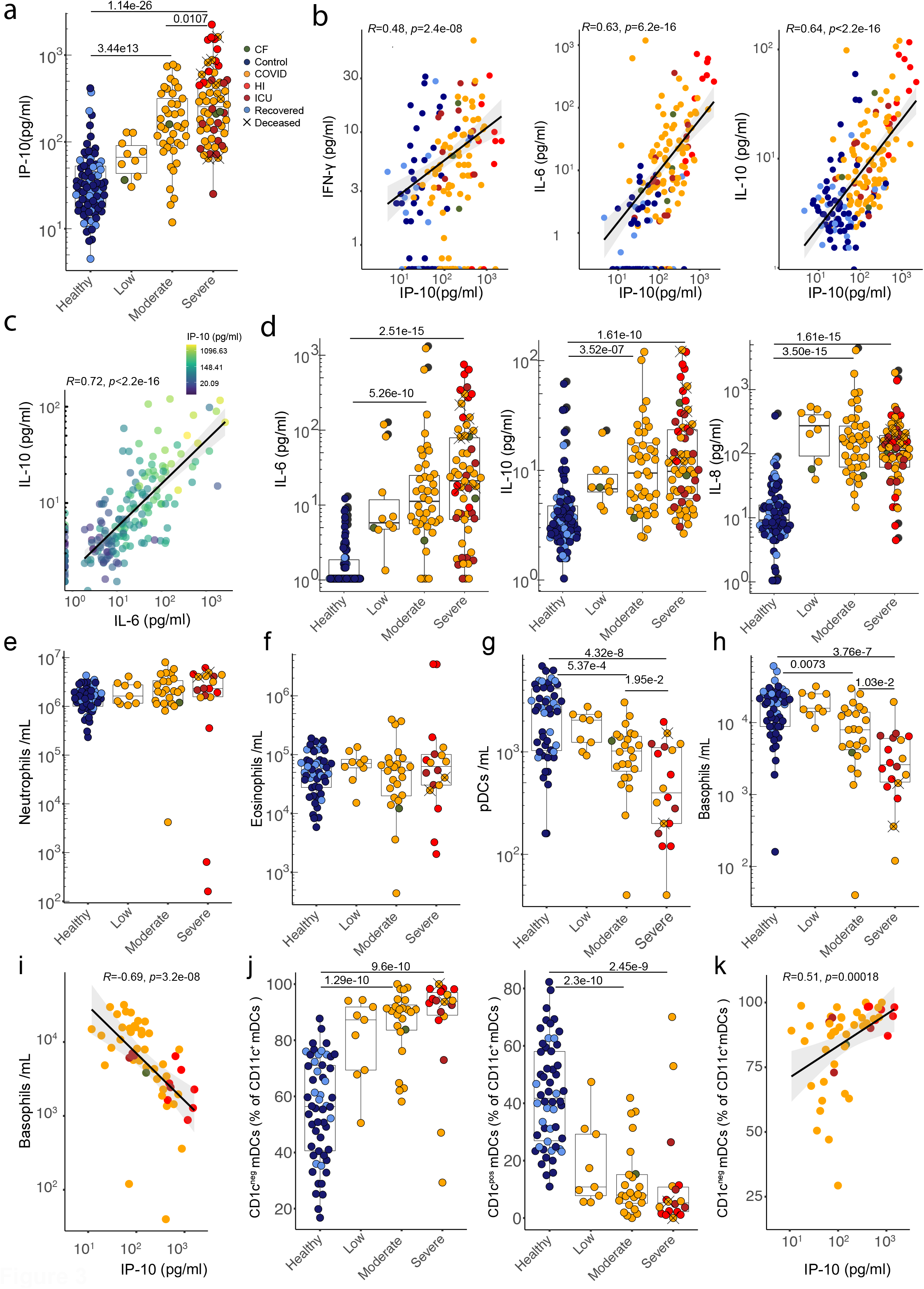
COVID-19 patients display dysregulated cytokine responses and numeric alterations in the myeloid compartment. **a)** Plasma IP-10 concentration. Correlations between plasma concentrations of **b)** IP-10 and IFN-γ, IL-6 and IL-10 and **c)** between IL-10 and IL-6, coloured by IP-10 concentration. **d)** Plasma concentrations of IL-6, IL-10 and IL-8. a-d) Healthy n=62 n’ =43, low n=10 n’ =6, moderate n=40 n’ =23, severe n=57 n’ =29. Quantification of **e)** neutrophils, **f)** eosinophils, **g)** plasmacytoid dendritic cells and **h)** basophils from whole blood. **e-h)** Healthy n=54 n’ =37, low n=9 n’ =5, moderate n=24 n’ =18, severe n=18 n’ =9. **i)** Correlation between plasma IP-10 concentration and basophil numbers in COVID samples. n=51 n’=22. **j)** Frequencies of CD1c^neg^ and CD1c^pos^ mDCs. Healthy n=54 n’ =37, low n=9 n’ =5, moderate n=24 n’ =18, severe n=18 n’ =9. **k)** Correlation between plasma IP-10 concentration and frequency of CD1c^neg^ mDCs in COVID patients. n=51 n’=22. n=samples, n’=individuals. n/n’ may vary slightly between graphs due to data filtering or experimental dropouts (see methods). Correlation plots display results from Spearman correlation tests and a linear regression line with 95% confidence interval shading. Box plots denote median and 25^th^ to 75^th^ percentiles (boxes) and 10^th^ to 90^th^ percentiles (whiskers) and were statistically evaluated by a linear mixed model grouped by severity (excluding low), with patient as random variable, corrected for age- and sex-dependency. CF: confounding factors; HI: hyperinflammatory; ICU: intensive care unit.

We hypothesised that high sustained IL8, IL6, and IP10 levels might be reflected in the cellular composition of a Covid-IP immune signature. However, whereas neutrophils are commonly increased in sepsis, and reportedly expanded in Covid-19^31,37^, the trend toward higher neutrophil counts in CP was not significant, despite widespread upregulation of IL8 that activates neutrophils (**Fig 3e**). Likewise, eosinophil counts were comparable across HC and CP (**Fig 3f**).

By contrast, there were dramatic, severity-related depletions of plasmacytoid dendritic cells (pDC), which might contribute to defective Type-I IFN responses reported in Covid-19^25,38^, and of basophils that have been implicated in tissue repair^39^ and in suppression of coagulation^40^, possibly germane to frequent thromboses in severe Covid-19 (**Fig 3g,h**). Neither pDC nor basophil depletion have been widely reported for sepsis^41,42^. Note that potentially confounding factors such as asthma, allergy or antihistamines could not account for basophil losses, that were instead strongly correlated inversely with IP10 (**Fig 3i**). Also across CP, blood DC composition changed significantly with the predominant CD11c^+^CD1c^neg^ subset showing active cell cycling and increased frequencies that also correlated strongly with IP10, and somewhat with IL6 although that was not significant (**Fig 3j,k; Supp Fig 3a-c**). In sum, whereas sustained levels of IL6, IL10, and IL8 were common in CP, increased IP10 levels contributed to a consensus signature, correlating with several other innate immune traits, that each correlated quantitatively and often prognostically with increased disease severity.

### Sepsis-like innate immune cell dysregulation

To investigate further the interplay of innate immune cells and cytokines in CP blood, we examined monocytes. As has been reported for healthy individuals^43^, the CV for HC monocytes was high, but a tendency of reduced numbers was still evident in CP (**Fig 4a**). Monocyte composition also changed, particularly with increased representation of CD16^+^CD14^+^ intermediate monocytes (MO^IM^) (**Fig 4a; Supp Fig 3d-f**), as was reported for responses to virus infection^44,45^. Nonetheless, the most overt phenotype was near-universal diminution of monocyte CD86 and HLA-DR that was most extreme in the severe sub-cohort (**Fig 4b**). Strikingly, residual CD1c^+^ DC (above) also showed decreased, severity-related HLA-DR expression (**Fig 4c**).

**Figure 4:**
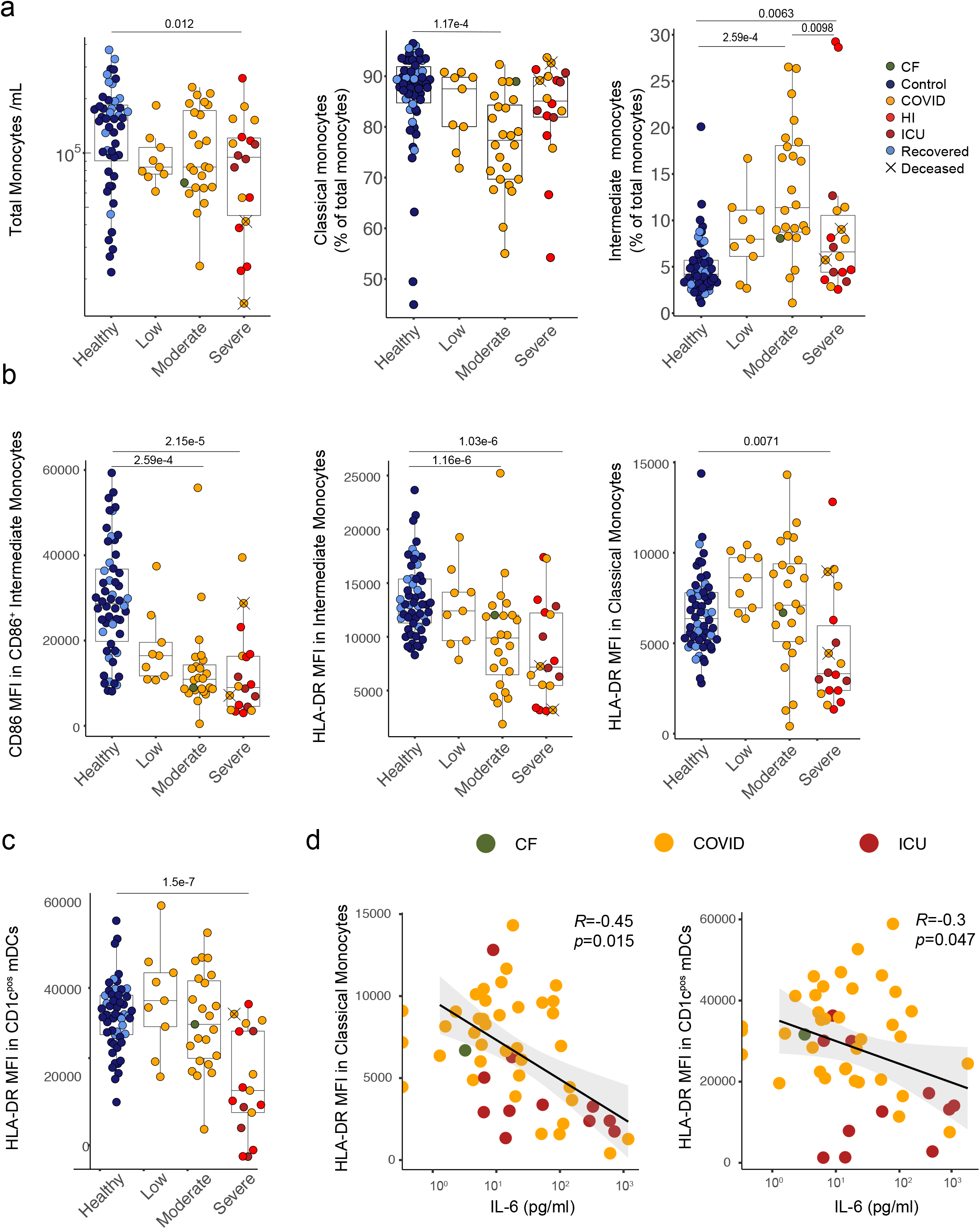
COVID-19 patients display a disrupted monocyte and dendritic cell phenotype. **a)** Quantification of monocyte number and frequencies of classical and intermediate monocytes in whole blood. **b)** MFI of CD86 in CD86^+^ intermediate monocytes and HLA-DR in intermediate and classical monocytes. **c)** MFI of HLA-DR in CD1c^pos^ mDCs. **d)** Correlation between plasma IL-6 concentration and HLA-DR MFI in classical monocytes and CD1c^pos^ mDCs. **a-d)** Healthy n=54 n’ =46, low n=9 n’ =5, moderate n=24 n’ =15, severe n=18 n’ =9. n=samples, n’=individuals. n/n’ may vary slightly between graphs due to data filtering or experimental dropouts (see methods). Correlation plots display results from Spearman correlation tests and a linear regression line with 95% confidence interval shading. Box plots denote median and 25^th^ to 75^th^ percentiles (boxes) and 10^th^ to 90^th^ percentiles (whiskers) and were statistically evaluated by a linear mixed model grouped by severity (excluding low), with patient as random variable, corrected for age- and sex-dependency. CF: confounding factors; HI: hyperinflammatory; ICU: intensive care unit.

CD86 and HLA-DR facilitate antigen-presentation by monocytes and DC, and it was conceivable that their low expression reflected cellular immaturity, in turn reflecting disease-associated myelopoiesis. This notwithstanding, suppressed HLA-DR expression is a marker of sepsis severity, and can be directly mediated by IL6 and by IL10^46^. Indeed, whereas there were many cases of low HLA-DR expression in the absence of IL6 overexpression, all but one patient with high IL6 displayed low HLA-DR expression on monocytes and/or DC (**Fig 4d**). In short, the consensus Covid-19 immunophenotype extended from strong associations of discrete cytokines and innate immune cell composition to sepsis-like states of atypical activation of monocytes and DC.

### Coexistent suppression and activation of T lymphocytes

Although pan-lymphopenia has been frequently cited in severe covid-19^37^, we observed primarily T cytopenia across all disease categories (**Fig 5a; Supp 4a,b**). NK cell depletion was significant but much less pronounced, akin to B cells (above) (**Fig 5a**). There were decreases in CD4^+^ and particularly CD8^+^ T cells, with consequent albeit variable increases in CD4:CD8 ratios (**Fig 5b**), and changes in the relative frequencies of many T cell subsets (**Supp Fig 4a**). γδ T cells were very severely depleted across all disease categories reflecting nearly ablated Vγ9Vδ2 cells that ordinarily dominate blood γδ cells (**Fig 5c, Supp Fig4c**). Some compensatory increases occurred in Vδ1^+^ cells, that are commonly tissue-associated, and which displayed many more cells in G_1_ compared to HC wherein almost all were G_0_ (see below). Conversely, NK cell composition, demarcated by CD56 and CD16 expression, seemed largely unaltered, particularly in moderate and severe disease (**Supp Fig4d**).

**Figure 5:**
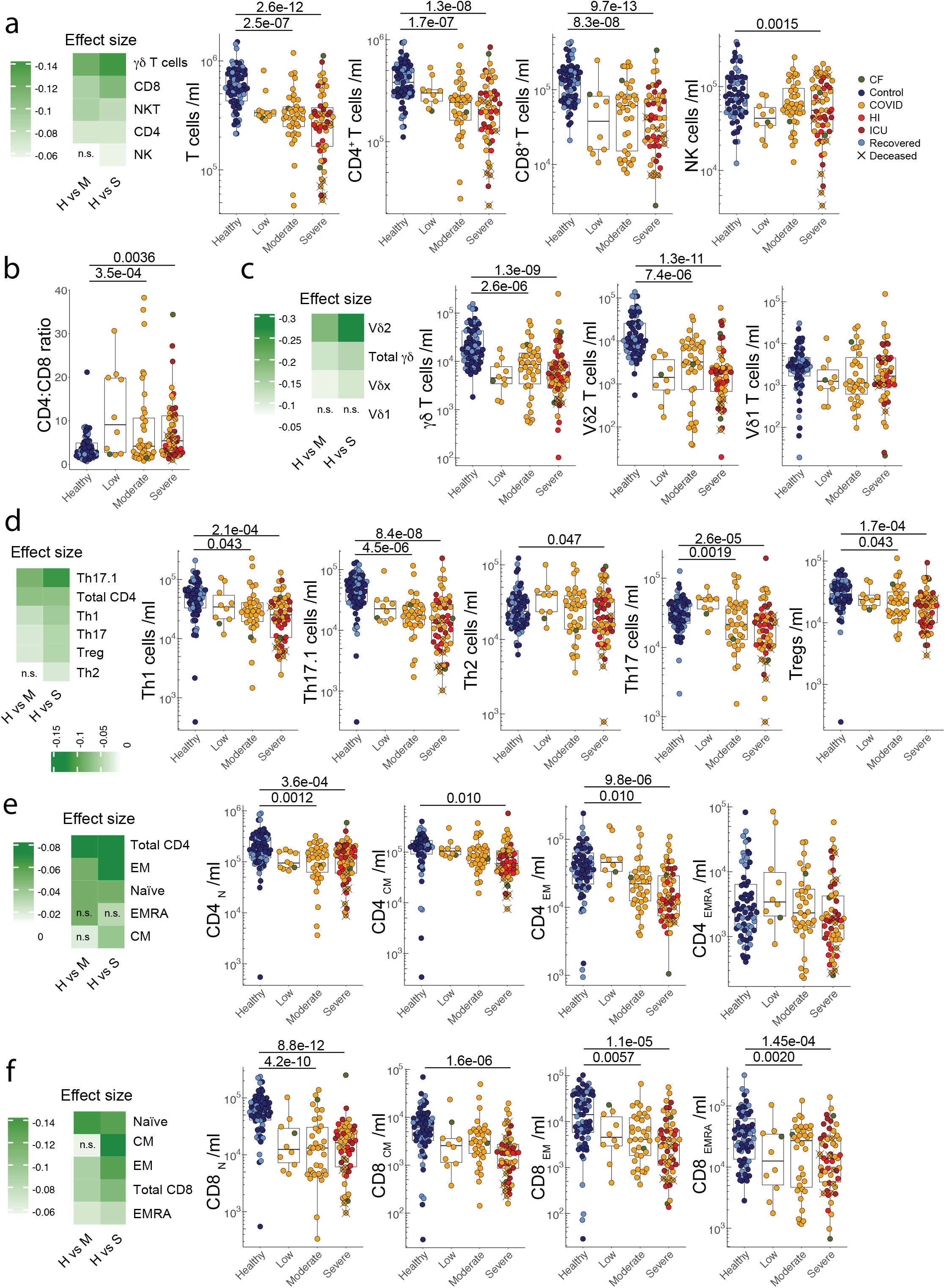
COVID patients display selective cytopenia in particular T cell subsets. **a)** Cytopenia effect size and quantification of total T cells, CD4, CD8 and NK cells. **b)** CD4 to CD8 ratio. Cytopenia effect size and quantification of **c)** total γδ T cells and subsets by Vδ usage, **d)** CD4 effector subsets, **e)** CD4 memory subsets and **f)** CD8 memory subsets. **a-f)** Healthy n=67 n’=51, low n=10 n’=6, moderate n=39 n’=24, severe n=58 n’=30. n=samples, n’=individuals. n/n’ may vary slightly between graphs due to data filtering or experimental dropouts (see methods). Box plots denote median and 25^th^ to 75^th^ percentiles (boxes) and 10^th^ to 90^th^ percentiles (whiskers) and were statistically evaluated by a linear mixed model grouped by severity (excluding low), with patient as random variable, corrected for age- and sex-dependency. Effect size is expressed as the difference in mean values divided by the Healthy mean value. CF: confounding factors; HI: hyperinflammatory; ICU: intensive care unit.

Strikingly, CP recapitulated the highly subset-specific T cytopenia pattern of sepsis. Thus, classifying Th subsets by chemokine receptor expression revealed substantial depletion, particularly in severe patients, of CD4^+^ Th_17.1_ cells and to some extent Th_1_ cells, which both produce IFNγ. As observed in sepsis, (Ghnewa et al., 2020) Th_2_ cells were much less affected, with intermediate impacts on Th_17_ and T_reg_ cells (**Fig 5d**). Cytopenia affected CD4^+^ and CD8^+^ T_EM_ and central memory (T_CM_) cells, but was not obviously limited to activated T cells, affecting naïve (T_N_) CD4^+^ cells, and being almost invariable for CD8^+^ T_N_ cells (**Fig 5e,f**). Putatively terminally differentiated CD8^+^ T_EMRA_ cells were significantly depleted across CP, but CD4^+^ T_EMRA_ were not, possibly because they were sustained by ongoing differentiation of activated T_EM_ cells (**Fig 5e,f**).

There were modest increases in the percentages of residual T cells expressing the activation marker, CD25 (IL2Rα), but overt increases in the frequency of activated HLA-DR^+^CD38^+^ cells, particularly among CD8^+^ T cells in patients progressing to severe disease who also showed the greatest CD8^+^ T_EM_cell depletions (above) (**Fig 6a; Supp Fig 5a**). Consistent with these overall findings, CD4^+^ T_EM_ cells purified from three HC subjects and from 3 CP patients with actively cycling T cells (see below), showed clear segregation of gene expression for *HLA-DR* and CD38, and for the cell-cycling associated protein detected with the Ki67 antibody (**Fig 6b**).

**Figure 6:**
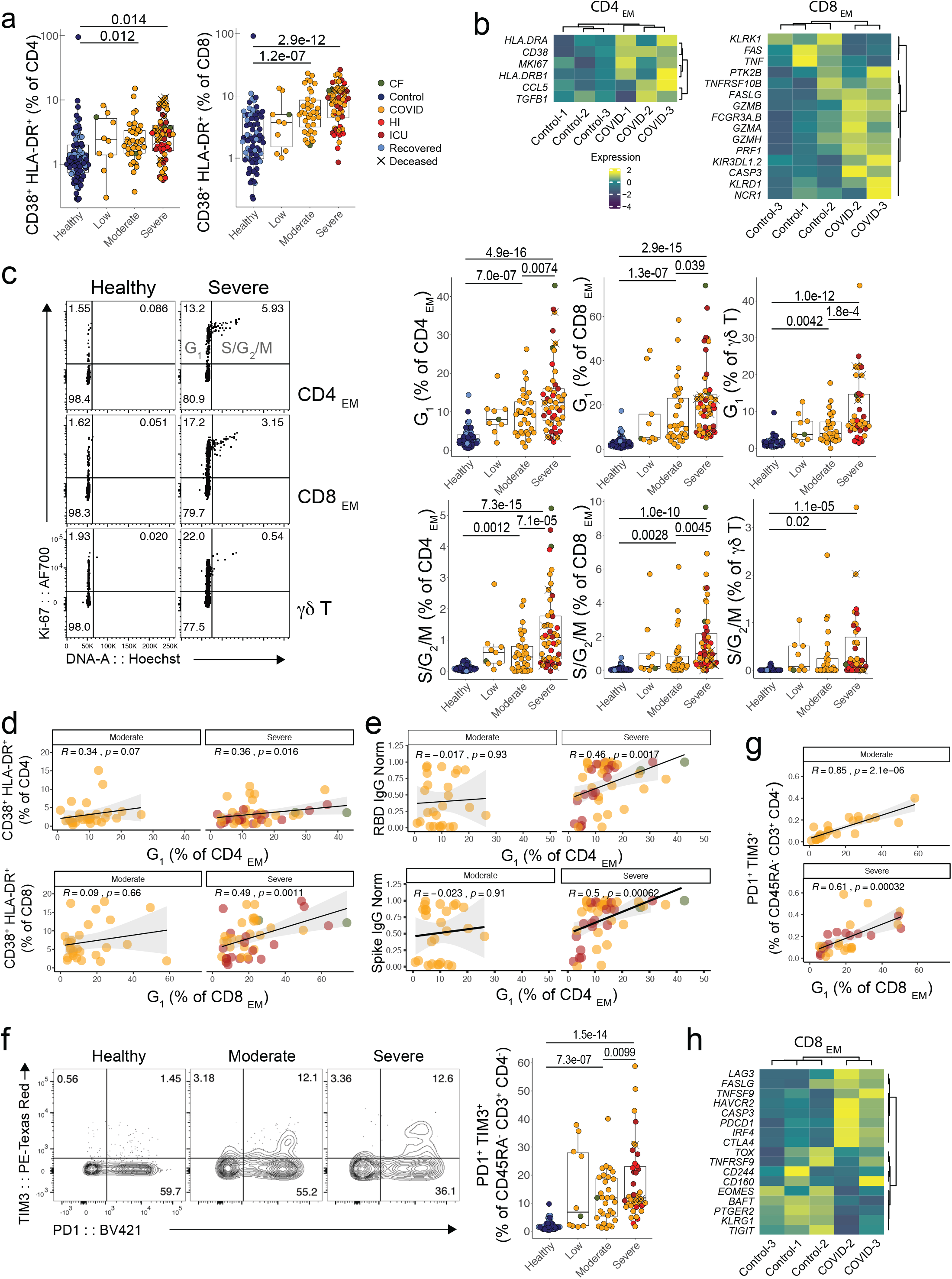
Co-existent activation and exhaustion of residual T cells in COVID patients. **a)** Frequency of activated (CD38^+^ HLA-DR^+^) CD4 and CD8 T cells. Healthy n=75 n’=55, low n=10 n’=6, moderate n=37 n’=22, severe n=56 n’=30. **b)** Hierarchical clustering of activation- and effector-associated gene expression in CD4 and CD8 effector memory cells from Nanostring analysis. Control n’=3, COVID n’=3 (CD4) or 2 (CD8). **c)** Representative flow cytometry and quantification of cell cycle status of CD4 and CD8 effector memory cells and γδ T cells. Healthy n=69 n’=53, low n=9 n’=5, moderate n=31 n’=20, severe n=48 n’=27. **d)** Correlations between activation and effector memory G1 frequencies in CD4 and CD8 T cells in moderate and severe COVID patients. **e)** Correlations between CD4 effector memory G1 frequency and anti-SARS-Cov-2 antibody titres. **d-e)** Moderate n=31 n’=20, severe n=48 n’=27. **f)** Representative flow cytometry and quantification of PD1^+^ TIM3^+^ effector memory CD8 T cell frequency. Healthy n=60 n’=50, low n=10 n’=6, moderate n=31 n’=19, severe n=38 n’=21. **g)** Correlation between PD1^+^ TIM3^+^ effector memory CD8 T cells and CD8 effector memory G1 frequencies in moderate and severe COVID patients. Moderate n=31 n’=19, severe n=38 n’=21. **h)** Hierarchical clustering of exhaustion- and effector-associated gene expression in CD8 effector memory cells from Nanostring analysis. Control n’=3, COVID n’=2. n=samples, n’=individuals. n/n’ may vary slightly between graphs due to data filtering or experimental dropouts (see methods). Correlation plots display results from Spearman correlation tests and a linear regression line with 95% confidence interval shading. Box plots denote median and 25^th^ to 75^th^ percentiles (boxes) and 10^th^ to 90^th^ percentiles (whiskers) and were statistically evaluated by a linear mixed model grouped by severity (excluding low), with patient as random variable, corrected for age- and sex-dependency. CF: confounding factors; HI: hyperinflammatory; ICU: intensive care unit.

Similarly, CD8^+^ T_EM_ cells, albeit from only 2 of the 3 CP patients, were clearly discriminated from HC CD8^+^ T_EM_ cells by genes suggesting strong, cytolytic effector potentials. Thus, whereas *KLRK1* (encoding NKG2D) and *TNF* were reduced, *NCR1* (encoding NKp46), *FASLG*, several *GZM* genes and PRF1 (encoding perforin) were all over-expressed relative to HC (Fig 6b). Additionally, overexpression of *TNFRSF10B* (encoding the TRAIL-receptor) and CASP3 (encoding caspase 3) suggested that CP CD8^+^ T_EM_ were more prone to apoptosis than HC T_EM_ (**Fig 6b**). By contrast, neither CD4^+^ nor CD8^+^ T_EM_ cells segregated from HC T_EM_ by expression of chemokine receptors that might have indicated a propensity to home to lungs or other tissues (**Supp Fig 5b**). However, CP CD4^+^ T_EM_ did show striking downregulation of genes encoding TCR signalling components and overexpression of HIF1 suggesting an adaptation to hypoxia or dysoxia, not seen in HC and strikingly evocative of sepsis (**Supp Fig 5c**).

Consistent with their activation, there were ∼10-fold increases in the percentage of blood CD4^+^ and CD8^+^ T cells in either G_1_ or S-G_2_/M phases of the cycle (so-called T_DS_ cells^32^ by contrast to HC in which ≥98% of cells comprise G_0_ cells in transit. These changes, illustrated for T_EM_ cells (**Fig 6c**), were strongly severity-related, and applied to all states of CD4^+^ and CD8^+^ cells, excepting T_N_ cells of which ≥99% remained in G_0_ in almost all patients (Supp Fig 5d). Likewise, there were ≥10-fold increases of residual γδ cells (mostly Vδ1^+^) in G_1_, albeit very few transitioned into S-G_2_ /M (**Fig 6c**). Unsurprisingly, the fractions of CD4^+^ and CD8^+^ T cells in G_1_ or S-G_2_ /M mostly correlated with the fraction of CD4^+^ and CD8^+^ cells expressing activation markers (**Fig 6d, Supp Fig 5e**, arrowed). Given that cues for T cell activation and cycling are received in the tissues and draining lymph nodes, assessing cell cycling in this fashion offers a systemic portal onto local immune dynamics.

The functional implications of activated CD4^+^ T cells were evident, for example, in significant correlations in severe patients of CD4^+^ T cells in G_1_ with RBD-specific and S-specific IgG (**Fig 6e**). Nonetheless, there was also a strong correlation of CD8^+^ T cells in G_1_ with CD8^+^ T cells co-expressing terminally differentiated/exhaustion markers, particularly PD1 and TIM3, which were almost undetectable in HC but which emerged in CP in a severity-related fashion (**Fig 6f,g, Supp Fig 5e**, arrowed), seemingly in parallel with CD4^+^ and CD8^+^ T cells in G_1_ or S-G_2_/M (**Fig 6c**, above). Furthermore, this chronic activation signature was evident in the increased expression in CP CD8^+^ T_EM_ cells *versus* HC T_EM_ cells of genes *PDCD1* (encoding PD-1), *LAG3*, and *CTLA4* (Fig 6h), although it was highly selective in that there was downregulation of another inhibitory receptor, TIGIT, whose ligand PVR may directly interact with SARS-CoV-2 proteins^47^.

### Anticipating severe disease progression

In sum, CP peripheral blood T cells displayed overt co-existence of activation, proliferation, effector differentiation, exhaustion and depletion. As in sepsis, the traits were selectively penetrant in different cell subsets. Were T cells to be correlates of protection, their dysregulation and depletion could be catastrophic, in which regard discrete T cell parameters commonly correlated prognostically to the severity of disease progression, akin to pDC and basophil depletion and elevated IP10. Given the intense interest in early, risk-based patient stratification, we next examined whether immunological parameters might discriminate hyper-inflammatory from other ICU patients, albeit in a small sample. Among four commonly-used clinical biomarkers, D-dimers and CRP were elevated in all patients and showed little discrimination among them; elevated procalcitonin associated only with HI; while highly elevated ferritin was mostly but not exclusively associated with HI (**Supp Fig 6a-d**). By contrast, highly elevated IP10 almost completely segregated HI patients from the others at each time-point examined (**Supp Fig 6e,f**; see also Fig 3A, above). Hence, whereas IP10 and ferritin levels correlate (**Supp Fig 6g**), IP10 measurements may have enhanced practical capacity to anticipate severe disease progression in the context of Covid-19.

## Discussion

Despite heterogeneity in gender, age, ethnicity, underlying condition, and manifestation of Covid-19, and despite some temporal variation in their sampling, the 63-strong CP cohort revealed a consensus, peripheral blood immune signature that combined elements of host-protective responses to virus infection with discrete elements of the immune landscape of sepsis; particularly atypical monocyte activation and T cytopenia. Those traits were applicable to the great majority of patients, as were IL8 overexpression, elevated plasmablasts, production of SARS-CoV-2-specific antibodies, reduced CD5^+^ B cell frequencies, and near-ablation of Vγ9Vδ2^+^ cells. Moreover, recent reports have variably described many of these traits in other diverse Covid-19 cohorts^6,23,27,37,45^, supporting the validity of a core Covid-19 immune signature, superimposed upon which may be more variable traits, such as neutrophil and NK cell numbers that were relatively unaffected in CP.

Further interrogating the Covid-19 immune signature, we identified discrete components that at Day 1 correlated, often prognostically, with the severity of disease progression, and frequently with one another. They include blood pDC and basophil depletion, increased IP10, and active cycling, activation and depletion of CD8^+^ T_EM_ cells. Hence, specific immune traits reflect and/or contribute to Covid-19 pathology. Indeed, pDC depletion may result in deficiencies in the Type-I IFN response reported for SARS-CoV-2 infection^25^. The collective prognostic potential of selected parameters might conceivably be engineered into a routine test in combination with other metrics, e.g. CRP and D-dimer, to facilitate risk-based patient stratification. Indeed, highly elevated IP10 levels showed better prognostic association than ferritin with progression to an HI state, albeit in a small sample. By contrast, all components of the Covid-IP signature, with the exception of anti-SARS-CoV-2 antibodies, scored as “normal” in HC subjects who had recovered from prior SARS-Cov-2 exposure without requiring hospitalisation, and IP10 was not elevated in several hospitalised patients suffering severe, non-Covid lower respiratory tract infections (LRTI) (AD, DD, and ACH; unpublished). Thus, an immuno-prognostic Covid-19 test might have high specificity.

Immunological correlates of protection were challenging to discern because of difficulties in quantifying virus loads, particularly in potentially inaccessible lower airway reservoirs. Although three patients who died failed to make SARS-Cov-2-specific antibodies, only one survived for long enough to mount such responses and he suffered from COPD. Moreover, no other correlations of Covid-IP severity with SARS-Cov-2-specific antibodies were evident, whereas severity correlated well with selective T cell proliferation, exhaustion and depletion. Correlations of cycling CD4^+^ T cells with anti-SARS-CoV-2 IgG, and the seemingly primed cytolytic status of CP CD8^+^ T_EM_ relative to HC CD8^+^ T_EM_ both strongly imply host-protective potentials of activated T cells.

T cells as key determinants of protection, might explain increased Covid-19 susceptibilities of older persons whose thymic involution depletes their potential to generate new T cell repertoires that are, by contrast, abundant in children. Moreover, adults’ greater reliance on memory T cells may be undermined by the signature dysregulation of T_EM_ cells in Covid-19. For example, the activated/proliferative phenotype for unprecedentedly high proportions of memory T cells in most CP patients might override and/or out-compete SARS-CoV-2-specific T cells. The cause of overt, subset-specific T cell dysregulation is unknown, and should be investigated with urgency. There was no evidence that CP T_EM_ expressed a different homing profile to HC T_EM_, but they segregated by gene expression related to apoptosis. Nonetheless, activation-induced cell death is challenging to reconcile with striking CD8^+^ T_N_ and CD4^+^ T_N_ depletions. Possibly, the expression by CD8^+^ T_EM_ of FasL, granzymes and perforin reflects fratricidal capabilities. Alternatively, different metabolic profiles of discrete T cell subsets might make them differentially susceptible to hypoxia, dysoxia, or specific inflammatory mediators.

Equally important insights into pathogenesis may emanate from the dysregulation of IP10 and of innate immune cells. Basophil depletion might reflect their recruitment into damaged lungs, thereby providing a surrogate marker of severity. However, depletion frequently preceded clinical evidence of severity, added to which basophils have been implicated in tissue repair, and in producing anti-coagulants; thus, their depletion might contribute to sustained pneumonitis and to thromboses commonly associated with Covid-19. Again, the drivers of basophil depletion are unelucidated. Near-universal IP10 upregulation is particularly interesting given the depletion of pDC, Th1, and Th17.1 cells that might portend deficiencies in Type-I and Type-II IFNs that are the main inducers of IP10. Possibly its levels are sustained by other, virus-related mechanisms. Elevated IP10 characterised SARS^48^ and MERS^38,49^, but not other sustained LRTI (see above). Moreover, MERS-CoV enters T cells *via* CD26, an ectopeptidase (as is ACE2, the receptor for SARS-CoV-2^38^ that regulates IP10 levels and activity^50^. Conceivably, IP10 dysregulation is a core component of pathogenic coronavirus infections, possibly interfering with ordered immunocyte chemotaxis. Thus, clues to Covid-19 pathogenesis provided by Covid-IP may guide therapeutic strategies, which, coupled with refined methods for tracking disease progression might ameliorate disease to such an extent that the pressure for virus eradication is reduced.

## Data Availability

All data and detailed protocols are available on the Covid-IP website (www.immunophenotype.org)

## Acknowledgments

We thank the patients and blood donors who agreed to take part in this study, as well as the medical and research team at Guy’s and St. Thomas Trust hospitals. We thank Furqan Shah, Zena Collins, Stelios Papaioannou, Celine Trouillet, Sara Cipolat, Amy Day, for logistics; Matt Brown, Sheeba Irshad, Mike Malim, and Stuart Neil; the Flow Cytometry core at the NIHR Biomedical Research Centre (BRC) of Guy’s and St Thomas’ Hospital (GSTT) and King’s College London (KCL); Richard Ellis, Manuela Terranova Barberio, Rozalyn Yorke, Fernanda Kyle, Sam Acors, Kathryn Steeel, Oliver Hemmings, Violetta Muñoz and Jonathan Warren for technical support and advice. We thank Google for research credits, Basecamp for free unlimited usage of their platform and volunteers at Crowdfight COVID-19. The work was supported by: a Cancer ImmunoTherapy Accelerator award from CRUK (AH; IDMDB), a CRUK City of London Major Cancer Centre studentship (M.J.); the Wellcome Trust (AH; PV; IZ; VS; 106292/Z/14/Z); the Rosetrees Trust (AH), King’s Together Seed Fund (AH), Gamma Delta Therapeutics (RD, DD, SH), The John Black Charitable Foundation (AH), and the Francis Crick Institute, (AH, LM; FC) which receives core funding from Cancer Research UK (FC001093), the MRC (FC001093) and the Wellcome Trust (FC001093) (AH); NIHR Clinician Scientist Award CS-2016-16-011 (MSH); the NIHR BRC for the Infectious Diseases Biobank (JC, CM), and for support of Infection and Immunity (JE; RJ112/N027); EMBO ALTF 198-2018 (MMR); an Irvington Fellowship-Cancer Research Institute (DM); National Institute of Academic Anaesthesia BJA-RCOA PhD Fellowship WKR0-2018-0047 (MF), NIHR Fellowship (AD, YW); Clinical Immunology Research Fund for the King’s College Hospital Charitable Trust (MI); UK MRC (MR/N013700/2) (CG); EMBL core funding and the European Union’s Horizon 2020 Research and Innovation Programme under grant agreement #730879 (FG, JB, JM), and Estonian Research Council grant PRG377 (PP). The authors also acknowledge King’s College London, the Francis Crick Institute and the EBI for institutional support. The views expressed are those of the authors and not necessarily those of the NHS, the NIHR or the Department of Health and Social Care.

## Author contributions

Methodology, A.G.L., A.L., I.D.M.D.B., A.D., L.Monin., M.M.R.

investigation, A.G.L., I.D.M.D.B., M.F., L.Monin., M.M.R., D.R.M., T.S.H., I.F.Q., R.D., A.J., I.Z., P.V., M.G., J.F., S.G., J.N.E.C., S.R., E.B.B., P.P., K.K., L.H., M.I., K.J.D., J.S., C.G.

Data analysis, A.G.L., A.L., I.D.M.D.B., A.D., M.F., L.Monin., M.M.R., D.R.M., T.S.H., I.F.Q., S.K., M.J., D.D., A.J., I.Z. Y.W., K.J.D., A.C.H.

interpretation, A.G.L., A.L., I.D.M.D.B., A.D., M.F., L.Monin., M.M.R., D.R.M., I.F.Q., S.K., M.J., D.D., P.V., Y.W., V.S., F.C., F.D.R., M.S.H., A.C.H.

Data curation A.G.L., A.L., T.S.H., J.M., F.L.G., K.Babalola; Project administration, S.K., R.D., E.T., C.M., J.C.

Sample provision, M.J., D.D., Y.W., L.Martinez, B.M., K.Bisnauthsing., K.Brooks., S.A.J.; Writing, A.C.H., M.S.H.

Conceptualization and supervision, J.E, M.S.H., A.C.H.

## Materials and Methods

### Study design and recruitment

Between 25^th^ March 2020 and 14^th^ May 2020, 63 patients (28-88 years of age, median 61) with confirmed SARS-CoV-2 infection by viral PCR (n=62) or serology alone (n=1) were recruited to the COVID-IP study from hospitals within Guy’s St Thomas’ NHS Trust for an observational cohort study with serial peripheral blood immunophenotyping and analysis of clinical outcomes (**Supp Table 1**). Blood sampling was performed within 24 hours of recruitment and, thereafter, approximately at day 3 and day 9 post recruitment with two patients having additional sampling timepoints.

Patients above 18 years of age and with capacity to consent were approached for informed consent to serial blood sampling by the research nursing team if they met the criteria of a positive PCR result for SARS-CoV-2 or strong clinical suspicion of infection. For patients in the intensive care unit (ICU) lacking capacity, this was sought from their next of kin or treating physician under appropriate ethical approval. Informed consent was obtained retrospectively from these patients, where possible.

Thus, the cohort includes patients admitted to the wards and ICU with symptomatic SARS-CoV-2 infection (n=56), patients admitted for other conditions who acquired SARS-CoV-2 during their stay (n=3) and ambulatory dialysis patients (n=4) diagnosed on screening tests. The majority of patients (n=55) were recruited within 72 hours of a positive PCR result or initial clinical suspicion. Due to the prospective nature of our sampling, we were able to capture a heterogenous population of ICU patients recruited prior to admission (n=3), during admission (n= 4) and immediately after ICU discharge (n= 8). Patients recruited post-ICU were sampled upon first encounter within a general medical ward.

During this same period, 55 healthy adult volunteers (25–71 years of age, median 36) with no known current malignancy, serious infectious illness, organ transplant or autoimmune disease were recruited as a control cohort for similar serial peripheral blood immunophenotyping. Given the nationwide lockdown during the period of the study recruitment, healthy adult volunteers were in main part research or clinical staff employed at Guy’s St Thomas NHS Trust or King’s College London. Potential donors were approached by the clinical research staff within the COVID-IP team for informed consent on a voluntary basis. A number of the healthy control volunteers (n=14) had experienced prior SARS-CoV-2 infections for which hospitalization had not been required and constitute a cohort of fully recovered, previously mildly infected individuals.

The study protocol for patient recruitment and sampling, out of the intensive care setting, was approved by the committee of the Infectious Diseases Biobank of King’s College London with reference number COV-250320. The protocol for healthy volunteer recruitment and sampling was

similarly approved by the same committee as an amendment to an existing approval for healthy volunteer recruitment with reference number MJ1-031218b. Both approvals are granted under the terms of the Infectious Disease Biobank’s ethics permission (reference 19/SC/0232) granted by the South Central Hampshire B Research Ethics Committee in 2019. Patient recruitment from the intensive care unit was undertaken through the ethics for the IMMERSE study approved by the South Central Berkshire Ethics Committee with reference number 19/SC/0187. Patient and control samples and data were anonymized at the point of sample collection by research nursing staff or clinicians involved in the COVID-IP project. We have complied with all relevant ethical regulations.

### Sample processing and cell isolation

Unfixed patient samples were handled under Biosafety Level 3 (BSL-3) containment conditions following risk assessments and code of practice approved by King’s College London. Blood samples in serum separator tubes were centrifuged at 1500 x g for 10min and serum aliquoted and stored at -80°C. Aliquots of blood from heparin tubes were stained for whole blood flow cytometry panels (see flow cytometry) or centrifuged at 2000 x g for 10min and plasma stored at -80°C. Remaining heparinised blood was diluted with 50% volume PBS, layered over Ficoll (GE Healthcare) in Leucosep tubes (Greiner Bio-One) and centrifuged at 800 x g for 15 min without break at room temperature. The PBMC fraction was then washed three times in cold PBS and used for flow cytometry. All flow samples were fixed for 10 min with either Cellfix (BD) or FoxP3 Fix/Perm kit (eBioscience) prior to removal from the BSL-3 facility.

### Flow Cytometry Staining and Acquisition

All flow cytometry antibodies and concentrations used for analysis can be found in **Supplementary Table 3**. PBMC samples were stained for viability with BD Horizon ™ Fixable Viability Stain 780. PBMC and whole blood cell surface staining was performed in BD Pharmingen ™ Stain Buffer (BSA) and BD Horizon Brilliant ™ Stain Buffer Plus for 30 min at room temperature. Intracellular staining was performed after permeabilizing cells with Invitrogen Permeabilization Buffer 10X for 30 min at 4°C. PBMC samples were stained using staining mix panels 1, 2, 3, 4, and 5 as found in **Supplementary Table 3**. 100μl of prepared PBMCs were spun down and resuspended in 100μl of staining mix. Cells were then washed in staining buffer and fixed for 10 minutes protected from light. Cells were then washed and resuspended in 200μl staining buffer for acquisition by flow cytometer for panels 1, 2, 3 and 4. PBMCs stained for panel 5 were resuspended in permeabilization buffer containing intracellular staining antibodies as found in **Supplementary Table 3**, and incubated at 4°C for 30 mins protected from light. The cells were then spun down and washed in DPBS before being resuspended in DPBS containing Hoechst 33342 (Thermofisher Scientific) and incubated at room temperature for 15 mins. Cells were washed, pelleted and resuspended in 200μl DPBS for acquisition by flow cytometer. Whole blood samples were stained using staining mix panels 6, 7 and 8 as found in Supplementary Table 3. 50μl of whole blood was stained in 50μl of antibody staining mix, washed in DPBS, and then fixed. Red blood cell lysis was then performed using eBioscience RBC Lysis Buffer (Multi-species) 10X diluted in deionized water for 15 min at room temperature. This was repeated up to two times to ensure adequate removal of red blood cells. Samples were then spun down and resuspended in 200μl staining buffer for acquisition by flow cytometer. For PBMC panels 1, 2, 3 and 4, 100μl of sample was analysed on a five laser BD LSRFortessa acquired with a BD High Throughput Sampler (HTS). For whole blood panels 6, 7 and 8, 100μl of sample was analysed on a 4 laser BD LSRFortessa acquired with a BD HTS. For PBMCs stained using panel 5, cells were acquired on a 4 laser BD LSRFortessa in FACS tubes, run on low for 10 mins, with samples diluted to achieve an event rate of no more than approximately 200 events/s.

### Cell Sorting by Flow Cytometry

PBMCs were stained with the following antibodies from the panels found in **Supplementary Table 3**; CD3-FITC (P5), TCR γδ-PE-Cy7 (P3), CD4-BV711 (P5), CD8-PerCP-Cy5.5 (P3), CD25-PE (P1: 2A3 & M-A251), CD45RA-BV786 (P3), CCR7-PE-CF594 (P3), and CD127-BV421 (Sort). TEM CD4 and TEM CD8⍰cells were gated using gating strategy (**Supp Fig 7**), and purified by FACS on a Becton Dickinson FACSAriaIII cell sorter equipped with four lasers. A 70-µm nozzle running at 70⍰p.s.i. and 90⍰kHz was used as the setup. FACS gates were drawn to include only live single cells based on BD Horizon™ Fixable Viability Stain 780.

### Flow Cytometry Data Analysis

FCS files were analysed using FlowJo (10.6.2, Treestar). Gating strategies for all panels are outlined in **Supplementary Figure 7**. Event counts for every gate were exported and frequencies to relevant parent populations were calculated in R. Absolute cell counts were back-calculated using the counts/mL of blood for major lineages derived from whole blood count panel (panel 6), where the equivalent of 25μl of whole blood was analysed per sample. Median fluorescent intensities were calculated using FlowJo for relevant markers on specific populations. For panels 1-4 and 6-7 a minimum threshold of 100 events per gate was used to investigate its subpopulations/measure MFI.

### Cytokine Analysis

The LegendPlex Human Anti-Virus Response panel (13-plex) (740390, Biolegend) and the LegendPlex Human Th Panel (13-plex) (740721, Biolegend) were used according to manufacturer’s instructions with some modifications. The assay was carried out in V-bottom 96-well plates. Plasma was thawed and diluted 2-fold with assay buffer before being tested. Mixed beads, detection antibodies and Streptavidin-PE were diluted 2-fold in assay buffer and 25μl of each reagent were used. Following 1.5 h incubation with mixed beads, wells were washed twice with wash buffer. Samples and standards were incubated with detection antibodies for 45 min. Next, Streptavidin-PE was added and plate was incubated for another 20 min. Finally, beads were washed once and resuspended in 200 μl wash buffer and acquired on a 4 laser BD LSR Fortessa X20. All incubation steps were carried out in the dark at room temperature, on an orbital shaker set at 600rpm. Data were analysed using the LegendPlex data analysis software Version 8 for Windows.

### NanoString gene expression analysis

The multiplexed NanoString nCounterTMCAR-T Characterization panel was used as expression assay for profiling 780 human genes (NanoString Technologies, Inc., Seattle, WA, USA). The assay was performed according to the manufacturer’s protocol. Pre-processing and normalization of the raw counts was performed with nSolver Analysis Software v4.0 (www.nanostring.com). Gene expression data were normalized by using the 10 housekeeping genes present in the CAR-T Characterization panel. The 6 spiked-in RNA Positive Control and the 8 Negative controls present in the panel were used to confirm the quality of the run.

### Serology Analysis: ELISA

N protein was obtained from Leo James and Jakub Luptak at LMB, Cambridge. The N protein used is a truncated construct of the SARS-CoV-2 N protein comprising residues 48-365 (both ordered domains with the native linker) with an N terminal uncleavable hexahistidine tag. N was expressed in **E. Coli** using autoinducing media for 7h at 37°C and purified using immobilised metal affinity chromatography (IMAC), size exclusion and heparin chromatography. S protein consists of pre-fusion S ectodomain residues 1-1138 with proline substitutions at amino acid positions 986 and 987, a GGGG substitution at the furin cleavage site (amino acids 682-685) and an N terminal T4 trimerisation domain followed by a Strep-tag II [18]. The plasmid was obtained from Philip Brouwer, Marit van Gils and Rogier Sanders at The University of Amsterdam. The protein was expressed in 1 L HEK-293F cells (Invitrogen) grown in suspension at a density of 1.5 million cells/mL. The culture was transfected with 325 µg of DNA using PEI-Max (1 mg/mL, Polysciences) at a 1:3 ratio. Supernatant was harvested after 7 days and purified using StrepTactinXT Superflow high capacity 50% suspension according to the manufacturer’s protocol by gravity flow (IBA Life Sciences). The RBD plasmid was obtained from Florian Krammer at Mount Sinai University [19]. Here the natural N-terminal signal peptide of S is fused to the RBD sequence (319 to 541) and joined to a C-terminal hexahistidine tag. This protein was expressed in 500 mL HEK-293F cells (Invitrogen) at a density of 1.5 million cells/mL. The culture was transfected with 1000 µg of DNA using PEI-Max (1 mg/mL, Polysciences) at a 1:3 ratio. Supernatant was harvested after 7 days and purified using Ni-NTA agarose beads.

All plasma samples were heat-inactivated at 56°C for 30 mins before use in the in-house ELISA. High-binding ELISA plates (Corning, 3690) were coated with antigen (N, S or RBD) at 3 µg/mL (25 µL per well) in PBS, either overnight at 4°C or 2 hr at 37°C. Wells were washed with PBS-T (PBS with 0.05% Tween-20) and then blocked with 100 µL 5% milk in PBS-T for 1 hr at room temperature. Wells were emptied and sera and plasma diluted at 1:50 and 1:25 respectively in milk were added and incubated for 2 hr at room temperature. Control reagents included CR3009 (2 µg/mL), CR3022 (0.2 µg/mL), negative control plasma (1:25 dilution), positive control plasma (1:50) and blank wells. Wells were washed with PBS-T. Secondary antibody was added and incubated for 1 hr at room temperature. IgM was detected using Goat-anti-human-IgM-HRP (1:1,000) (Sigma: A6907) and IgG was detected using Goat-anti-human-Fc-AP (1:1,000) (Jackson: 109-055-043-JIR). Wells were washed with PBS-T and either AP substrate (Sigma) was added and read at 405 nm (AP) or 1-step TMB substrate (Thermo Scientific) was added and quenched with 0.5 M H2S04 before reading at 450 nm (HRP). Data was normalised using a min/max normalisation in order to compare samples across batches. Values above 0.3 were considered positive.

### Serology Analysis LIPS assay

SARS-CoV-2 (NCBI Acc # NC_045512.2) RBD domain of S (aa 329-538) and N (aa 2-419) gene fragments were cloned into pNanoLuc vector, transfected into HEK293 cells, the cell lysates containing NanoLuc-fusion proteins were probed with plasma samples (0.5 - 11×110^6^ luminescence units; LU) for 1 hr at RT. The Protein G Sepharose beads (251µl of 4% suspension, Creative BioMart) were used to capture the immune complexes of anti-SARS-CoV-2 antibodies and NanoLuc fusion proteins. After washing, Nano-Glo™ luciferase substrate (Promega) was added and luminescence was measured in VICTOR X Multilabel Plate Readers (PerkinElmer Life Sciences). LIPS data represent the average of three replicative experiments. Results were given as fold changes (FC=LU sample/average LU of healthy control samples). A fold change greater than 2 was considered positive.

### Autoantibody screening

Thyroglobulin and thyroid peroxidase autoantibodies were measured in the serum by the electrochemiluminescence immunoassay (ECLIA) on the cobas analyser according to the recommendations of the manufacturer (Roche Diagnostics, Indianapolis, USA).

### Statistical Analysis

Cell subset counts (per mL of blood) and cytokine concentration were analysed after log10 transformation; all other parameters (cell subsets’ frequencies, serology parameters) were analysed without any additional data transformation.

Influence of age and sex was tested by comparing nested linear mixed models on healthy control data:

parameter∼1+(1|patient)

parameter∼1+age +(1|patient) or parameter∼1+sex +(1|patient)

parameter∼1+age+sex+ (1|patient)

For the cases when not enough samples were available to estimate patient effects, linear model was used. Correction with sex and/or age was used whenever a model with additional parameters was better according to Akaike criterion and with pval <0.01 (list of corrected parameters and estimated sex/age effect **(Supp Table 2**). For these parameters, values predicted based on age/sex influence estimates from healthy individuals were subtracted from the raw parameter values and residuals were used for downstream statistical testing.

Testing for difference between CP/controls and severity groups, raw values (or residuals, where sex/age was significant) between CP and controls were compared by fitting linear mixed model: parameter∼control_CPstatus+(1|patient) parameter∼severity+(1|patient), where severity was defined as Moderate for WHO 3-4 and Severe for WHO 5-8, Healthy for CP (Low were excluded in all comparisons except peak Ig titres). For the cases when not enough samples were available to estimate patient effects, linear model was used instead. Appropriate contrasts (moderate vs. healthy, severe vs. healthy, severe vs. moderate, CP vs controls) were extracted and effect size estimated by dividing difference between estimated means of populations by standard deviation of controls, unless stated otherwise (for age and sex corrected values this was done on age and sex corrected parameters).

Spearman correlations between parameters in the flow cytometry, serology and cytokine analysis were identified separately in Covid-19 and control cohorts (unless stated otherwise in the text). Correlations with *R* ≥ 0.3| *R*<-0.3 and *P* < 0.01 were considered significant.

### Data Availability

All data and detailed protocols are available on the Covid-IP website (www.immunophenotype.org).

---

**Supplementary Table 1.**
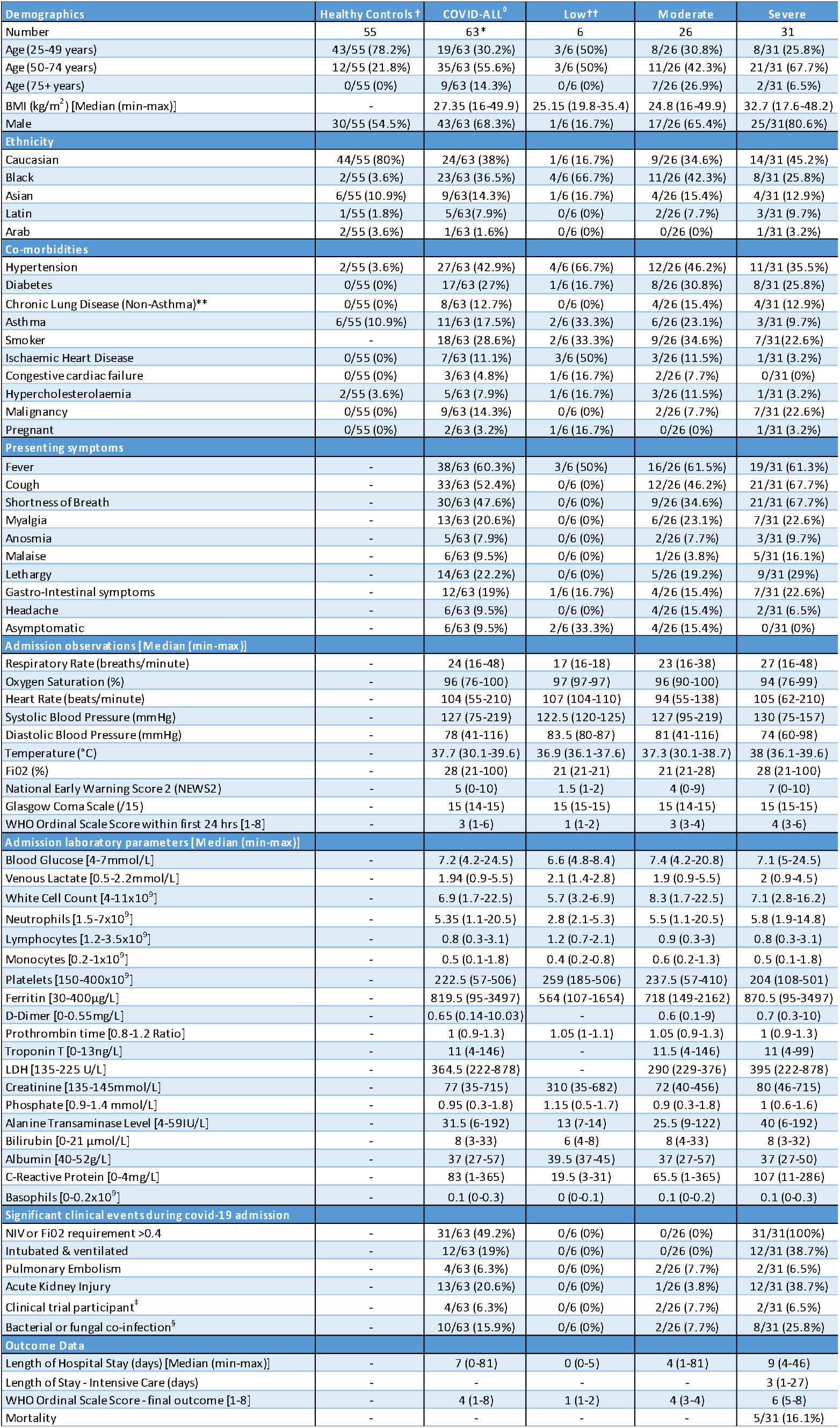
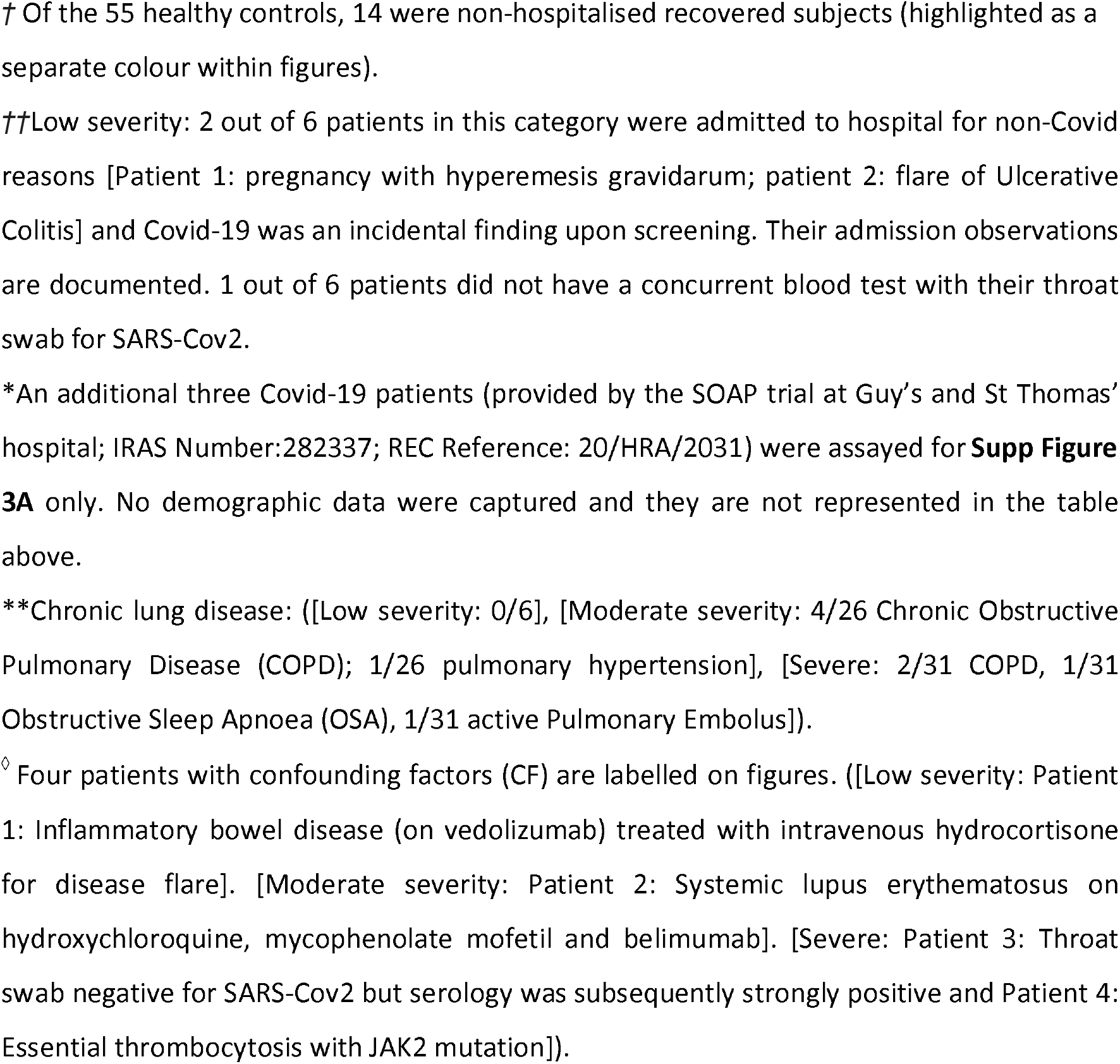
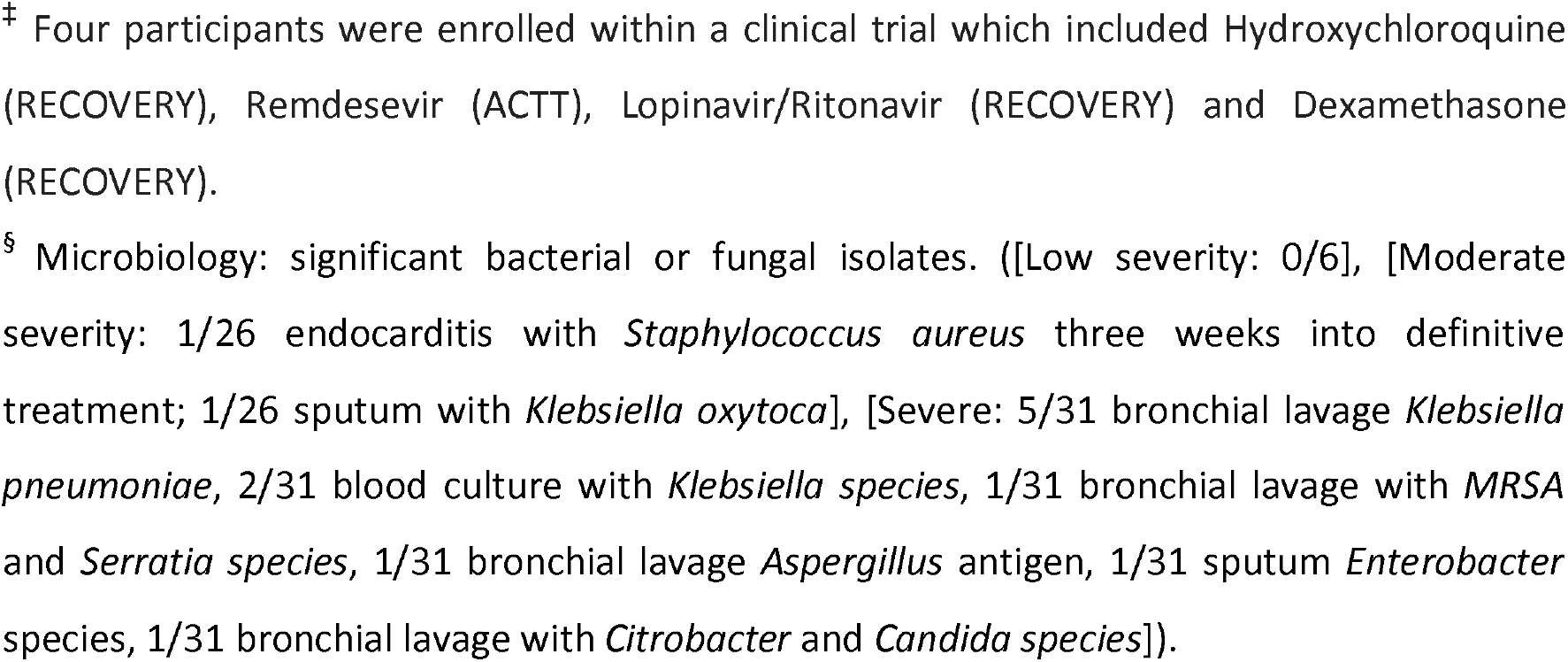
Characteristics of controls (n=55) and patients with Covid 19 (n=63). An eight-point ordinal scale was used to categorise Covid-19 patients by severity of disease; Low (1-2), Moderate (3-4) or Severe (5-8). The categories are: 1) No limitation of activities; 2) Limitation of activities; 3) Requiring hospitalisation – no oxygen therapy requirement; 4) Requiring oxygen therapy – *via* nasal cannula/Venturi mask (Fi02 <40%); 5) Requiring Non-Invasive Ventilation or high-flow oxygen therapy (Fi02 ≥40%); 6) Requiring intubation and mechanical ventilation; 7) Requiring mechanical ventilation and additional organ support; 8) Death.

**Supplementary Table 2:** Parameters for which statistical significance was altered by correction for age- and sex-dependency

| Parameter | Correction | Age effect estimate | Sex effect estimate | Age p-val | Sex p-val | Sig. change |
| --- | --- | --- | --- | --- | --- | --- |
| CD8_CD25p_01 \| Median (CD25) | age | 10.406 | NA | 0.001 | NA | — |
| CD8_CD25p_02 \| Median (CD25) | age | 8.031 | NA | 0.007 | NA | — |
| CD4_CD103p_03 \| Median (CD103) | sex | NA | 1644.309 | NA | 0.022 | — |
| B_01/CD45_01 \| freq | age | 0.001 | -0.019 | 0.036 | 0.070 | — |
| CD4_nonTreg_01/CD4_01 \| freq | sex | NA | -0.032 | NA | 0.001 | — |
| CD4_CCR4p_02/CD4_02 \| freq | age + sex | 0.002 | 0.031 | 0.005 | 0.029 | — |
| CD4_Th2_02/CD4_02 \| freq | age + sex | 0.002 | 0.014 | 0.000 | 0.028 | — |
| CD4_CD25p_02/CD4_02 \| freq | age | 0.003 | 0.047 | 0.016 | 0.145 | — |
| CD4_Treg_02/CD4_02 \| freq | sex | NA | 0.025 | NA | 0.001 | — |
| gd_Vd2_03/T_03 \| freq | age | -0.001 | NA | 0.023 | NA | — |
| CD19p_igMn_CD27p_igGp_4/B_4 \| freq | sex | -0.001 | -0.023 | 0.071 | 0.006 | — |
| PD1n_7/<br>CD4p_CD45RAn_CXCR3n_CCR6p_7<br>\| freq | age | 0.003 | -0.040 | 0.005 | 0.051 | + |
| PD1p_7/<br>CD4p_CD45RAn_CXCR3n_CCR6p_7<br>\| freq | age | -0.003 | 0.040 | 0.005 | 0.051 | + |
| CD3p_CD4n_PD1n_7 \| Count_back | sex | -0.011 | -0.333 | 0.081 | 0.026 | — |
| CD4n_CD45RAn_CXCR3p_CCR6n_7<br>\| Median (LAG3) | age | 1.885 | NA | 0.008 | NA | — |
| CD4n_CD45RAn_TIM3p_PD1n_7<br>\| Count_back | sex | NA | -0.321 | NA | 0.044 | — |
| CD4n_CD45RAp_PD1n_7 \| Count_back | age | -0.018 | -0.319 | 0.037 | 0.115 | — |
| CD4p_CD45RAn_CXCR3n_CCR6n_7<br>\| Count_back | age | 0.010 | NA | 0.000 | NA | + |
| CD4p_CD45RAn_CXCR3n_CCR6p_7<br>\| Count_back | age + sex | 0.010 | -0.214 | 0.028 | 0.048 | + |
| CD4p_CD45RAn_TIM3n_PD1p_7 \|<br>Count_back | age + sex | 0.007 | -0.207 | 0.049 | 0.024 | — |
| B_Cells_08 \| Median (CD86) | sex | NA | 110.875 | NA | 0.037 | + |
| mDC_08/mDC2_08 \| Count_back | age | 0.013 | NA | 0.010 | NA | — |
| CD19p_igMn_CD27p_igGp_4 \|<br>Count_back | age | -0.026 | NA | 0.016 | NA | — |
| Transitional_4/T2_4 \| Count_back | age | -0.033 | NA | 0.047 | NA | + |
| CD4_Th17_02 \| Count_back | age | 0.006 | NA | 0.027 | NA | + |

Age- and sex-correction was applied as detailed in methods. Count\_back parameters were analysed following log transformation. Statistical comparison between healthy, moderate and severe samples was performed as a linear mixed model grouped by severity, with patient as random variable, as detailed in methods. Parameters listed above either gained (+) or lost (-) statistical significance following age and/or sex correction as outlined.

**Supplementary Table 3:** Antibody panels for flow cytometery analysis

| Panel | Target | Fluorophore | Clone | Brand | Volume per 100 microlitres |
| --- | --- | --- | --- | --- | --- |
| 1 | CD103 | BV421 | BER-ACT08 | BD | 1.25 |
| 1 | CD127 | BV786 | HIL-7R-M21 | BD | 2.5 |
| 1 | CD14 | BV711 | MφP9 | BD | 0.63 |
| 1 | CD16 | PerCP-Cy5.5 | 3G8 | BD | 0.16 |
| 1 | CD19 | BUV737 | SJ25C1 | BD | 1.25 |
| 1 | CD25 | PE | 2A3 | BD | 5 |
| 1 | CD25 | PE | M-A251 | BD | 5 |
| 1 | CD27 | BV605 | L128 | BD | 1.25 |
| 1 | CD3 | BUV395 | UCHT1 | BD | 2.5 |
| 1 | CD4 | BV510 | SK3 | BD | 1.25 |
| 1 | CD45 | AF700 | HI30 | BD | 1.25 |
| 1 | CD45RA | PE-Cy7 | HI100 | BD | 1.25 |
| 1 | CD56 | PE-CF594 | NCAM16.2 | BD | 0.625 |
| 1 | CD8 | FITC | SK1 | BD | 2.5 |
| 1 | NKG2D | APC | 1D11 | BD | 10 |
| 2 | CCR4 | APC | 1G1 | BD | 2.5 |
| 2 | CCR6 | BV421 | 11A9 | BD | 2.5 |
| 2 | CCR7 | PE-CF594 | 150503 | BD | 5 |
| 2 | CD103 | BV711 | BER-ACT08 | BD | 2.5 |
| 2 | CD127 | BV786 | HIL-7R-M21 | BD | 2.5 |
| 2 | CD25 | PE | 2A3 | BD | 5 |
| 2 | CD25 | PE | M-A251 | BD | 5 |
| 2 | CD27 | BV605 | L128 | BD | 1.25 |
| 2 | CD3 | BUV395 | UCHT1 | BD | 2.5 |
| 2 | CD38 | BUV737 | HB7 | BD | 0.63 |
| 2 | CD4 | BV510 | SK3 | BD | 1.25 |
| 2 | CD45 | AF700 | HI30 | BD | 1.25 |
| 2 | CD45RA | PE-Cy7 | HI100 | BD | 1.25 |
| 2 | CD8 | FITC | SK1 | BD | 2.5 |
| 2 | CXCR3 | PE-Cy5 | 1C6 | BD | 10 |
| 2 | HLA-DR | PerCP-Cy5.5 | L243 | BD | 5 |
| 3 | CCR7 | PE-CF594 | 150503 | BD | 5 |
| 3 | CD103 | BV711 | BER-ACT08 | BD | 2.5 |
| 3 | CD27 | BV605 | L128 | BD | 1.25 |
| 3 | CD3 | BUV395 | UCHT1 | BD | 2.5 |
| 3 | CD4 | BV510 | SK3 | BD | 1.25 |
| 3 | CD45 | AF700 | HI30 | BD | 1.25 |
| 3 | CD45RA | BV786 | HI100 | BD | 1.25 |
| 3 | CD8 | PerCP-Cy5.5 | SK1 | BD | 1.25 |
| 3 | NKG2D | APC | 1D11 | BD | 10 |
| 3 | PD-1 | BV421 | EH12.1 | BD | 1.25 |
| 3 | TCR γδ | PE-Cy7 | IMMU510 | Beckman | 2.5 |
| 3 | Vδ1 | FITC | REA173 | Miltenyi | 0.5 |
| 3 | Vδ2 | PE | B6 | BD | 1.25 |
| 4 | CD19 | BV711 | SJ25C1 | BD | 2 |
| 4 | CD24 | BUV395 | ML5 | BD | 2 |
| 4 | CD25 | APC-R700 | 2A3 | BD | 2 |
| 4 | CD27 | BV786 | L128 | BD | 2 |
| 4 | CD38 | PE | HIT-2 | BD | 10 |
| 4 | CD43 | BV421 | 1610 | BD | 2 |
| 4 | CD5 | PE-Cy7 | L17F12 | BD | 2 |
| 4 | HLA-DR | BV510 | G46-6 | BD | 2 |
| 4 | IgD | BUV737 | IA6-2 | BD | 2 |
| 4 | IgG | APC | G18-145 | BD | 5 |
| 4 | IgM | BB515 | G20-127 | BD | 2 |
| 5 | CCR7 | PE-CF594 | 150503 | BD | 5 |
| 5 | CD25 | PE | 2A3 | BD | 5 |
| 5 | CD25 | PE | M-A251 | BD | 5 |
| 5 | CD3 | FITC | UCHT1 | BD | 10 |
| 5 | CD4 | BV711 | SK3 | BD | 1.25 |
| 5 | CD45RA | BV786 | HI100 | BD | 1.25 |
| 5 | CD8 | PerCP-Cy5.5 | SK1 | BD | 1.25 |
| 5 | TCR $\gamma\delta$ | PE-Cy7 | IMMU510 | Beckman | 2.5 |
| 5 | Ki-67 (intracellular) | AF700 | B56 | BD | 1 |
| 5 | FoxP3 (intracellular) | AF647 | 259D | Biolegend | 1 |
| 6 | CD10 | BV711 | HI10a | Biolegend | 2.5 |
| 6 | CD14 | AF488 | HCD14 | Biolegend | 1 |
| 6 | CD15 | BV605 | W6D3 | Biolegend | 0.25 |
| 6 | CD16 | PE-Cy7 | 3G8 | Biolegend | 0.125 |
| 6 | CD19 | PE | HIB19 | Biolegend | 0.5 |
| 6 | CD3 | APC-Cy7 | OKT3 | Biolegend | 0.5 |
| 6 | CD45 | PerCP | HI30 | Biolegend | 0.5 |
| 6 | CD56 | APC | HCD56 | Biolegend | 2 |
| 6 | CD62L | BV785 | DREG-56 | Biolegend | 1 |
| 6 | CD64 | BV421 | 10.1 | Biolegend | 2.5 |
| 7 | 2B4 | APC | (2-69) | BD | 2.5 |
| 7 | CCR6 | BB515 | 11A9 | BD | 2.5 |
| 7 | CD3 | APC-Cy7 | OKT3 | Biolegend | 1 |
| 7 | CD4 | PE-Cy7 | RM4-5 | BD | 0.5 |
| 7 | CD45RA | BV786 | HI100 | BD | 1 |
| 7 | CXCR3 | BB700 | CXCR3-173 | BD | 2.5 |
| 7 | LAG-3 | BV510 | TA7-530 | BD | 2.5 |
| 7 | PD-1 | BV421 | EH12.1 | BD | 2.5 |
| 7 | TIM3 | PE-CF594 | 7D3 | BD | 2.5 |
| 8 | CD11c | BV650 | 3.9 | Biolegend | 1 |
| 8 | CD123 | PerCP-Cy5.5 | 6H6 | Biolegend | 1 |
| 8 | CD14 | BV421 | M5E2 | Biolegend | 1 |
| 8 | CD16 | PE-Cy7 | 3G8 | Biolegend | 0.5 |
| 8 | CD19 | BV786 | HIB19 | Biolegend | 1 |
| 8 | CD1c | APC | L161 | Biolegend | 1 |
| 8 | CD3 | AF700 | OKT3 | Biolegend | 0.5 |
| 8 | CD303 | FITC | 201A | Biolegend | 1 |
| 8 | CD86 | PE | IT2.2 | Biolegend | 1 |
| 8 | HLA-DR | BV510 | G46-6 | BD | 1 |
| Sort | CD127 | BV421 | A019D5 | Biolegend | 1 |

## Figure Legends

**Supplementary Figure 1: Co-existent anti-SARS-Cov-2 antibody responses and disruption to the B cell compartment in COVID patients.**

Peak **a)** IgM and **b)** IgG titres against SARS-COV-2 N antigen. **a-b)** Healthy n’=48, Low n’=6, Moderate n’=25, Severe n’=31. **c)** Correlation between ELISA and LIPS serology measurements. COVID n = 76, n’ = 37. **d)** Log2 fold change of B cell frequency parameters between control and COVID cohorts. Control n=71 n’=52, COVID n=96 n’=57. **e)** WHO ordinal scale severity assessment of a cohort of COVID patients categorised by anti-thyroid autoantibody serology (patients with anti-thyroglobulin ≥115 IU/mL and/or anti-thyroid peroxidase ≥34 IU/mL were deemed positive). Ab negative n’ = 21, Ab positive n’ = 5. n=samples, n’=individuals. n/n’ may vary slightly between graphs due to data filtering or experimental dropouts (see methods). Correlation plots display results from Spearman correlation tests and a linear regression line with 95% confidence interval shading. Box plots denote median and 25^th^ to 75^th^ percentiles (boxes) and 10^th^ to 90^th^ percentiles (whiskers) and were statistically evaluated by a linear mixed model grouped by severity, with patient as random variable, corrected for age- and sex-dependency. CF: confounding factors; HI: hyperinflammatory; ICU: intensive care unit.

**Supplementary Figure 2: COVID-19 patients display dysregulated cytokine responses.**

**a)** Longitudinal measurement of plasma IP-10 concentration. Repeat samples from the same individual are linked. For healthy controls, longitudinal samples are those co-analysed with patients on the denoted days since admission. **b)** Plasma IFN-γ concentration. **a-b)** Healthy n=75 n’=55, low n=10 n’=6, moderate n=37 n’=22, severe n=56 n’=30. **c)** Heatmap of hierarchically clustered, quantile scaled plasma cytokine concentrations from all individuals. 22 unique cytokines measured. Control n=78 n’=55, COVID n=113 n’=63. n=samples, n’=individuals. n/n’ may vary slightly between graphs due to data filtering or experimental dropouts (see methods). Box plots denote median and 25^th^ to 75^th^ percentiles (boxes) and 10^th^ to 90^th^ percentiles (whiskers) and were statistically evaluated by a linear mixed model grouped by severity (excluding low), with patient as random variable, corrected for age- and sex-dependency. CF: confounding factors; HI: hyperinflammatory; ICU: intensive care unit.

**Supplementary figure 3: COVID-19 patients display a disrupted monocyte and dendritic cell phenotype.**

**a)** Representative flow cytometric cell cycle analysis of classical, intermediate and patrolling monocytes and CD1c^pos^ and CD1c^neg^ mDCs. **b)** Quantification of cell cycle analysis of CD1c^neg^ mDCs. Control n’=7, COVID n’=5. Unpaired Student’s t-test. Mean±SD. **c)** Correlation between plasma IL-6 concentration and CD1c^neg^ mDC frequency in COVID patients. n=51 n’=22. Log2 fold change in innate immune population **d)** frequency and **e)** absolute count parameters in COVID patients relative to controls. **d-e)** Control n=48 n’=34, COVID n=51 n’=22. **f)** Quantification of classical, intermediate and patrolling monocyte numbers from whole blood. Healthy n=48 n’ =34, low n=9 n’ =5, moderate n=24 n’ =16, severe n=18 n’ =9. n=samples, n’=individuals. n/n’ may vary slightly between graphs due to data filtering or experimental dropouts (see methods). Correlation plots display results from Spearman correlation tests and a linear regression line with 95% confidence interval shading. Box plots denote median and 25^th^ to 75^th^ percentiles (boxes) and 10^th^ to 90^th^ percentiles (whiskers) and were statistically evaluated by a linear mixed model grouped by severity (excluding low), with patient as random variable, corrected for age- and sex-dependency. CF: confounding factors; HI: hyperinflammatory; ICU: intensive care unit.

**Supplementary Figure 4: COVID patients display selective cytopenia in particular T cell subsets.**

Log 2 fold change of T cell **a)** frequency and **b)** absolute count parameters in COVID patients relative to controls. Healthy n=69 n’=53, COVID n=107 n’=60. **c)** Representative flow cytometry and quantification of γδ T cell subsets. **d)** Representative flow cytometry and quantification of NK cell subsets. **c-d)** Healthy n=69 n’=53, Low n=10 n’=6, Moderate n=39 n’=24, Severe n=58 n’=30. n=samples, n’=individuals. n/n’ may vary slightly between graphs due to data filtering or experimental dropouts (see methods). Box plots denote median and 25^th^ to 75^th^ percentiles (boxes) and 10^th^ to 90^th^ percentiles (whiskers) and were statistically evaluated by a linear mixed model grouped by severity (excluding low), with patient as random variable, corrected for age- and sex-dependency. CF: confounding factors; HI: hyperinflammatory; ICU: intensive care unit.

**Supplementary Figure 5: Co-existent activation and exhaustion of residual T cells in COVID patients.**

**a)** Frequency of CD25^+^ CD4 and CD8 T cells. Healthy n=75 n’=55, Low n=10 n’=6, Moderate n=37 n’=22, Severe n=56 n’=30. **b)** Hierarchical clustering of chemokine receptor gene expression in CD4 and CD8 effector memory cells by Nanostring analysis. **c)** Hierarchical clustering of TCR-dependent gene expression in CD4 effector memory cells by Nanostring analysis. **b-c)** Control n’=3, COVID n’=3 (CD4) or 2 (CD8). **d)** Flow cytometric cell cycle analysis of naïve CD4 and CD8 cells. Healthy n=67 n’=51, low n=10 n’=6, moderate n=39 n’=24, severe n=58 n’=30. **e)** Spearman correlations of all T cell frequency parameters in COVID patients. COVID n=114 n’=63. n=samples, n’=individuals. n/n’ may vary slightly between graphs due to data filtering or experimental dropouts (see methods). Box plots denote median and 25^th^ to 75^th^ percentiles (boxes) and 10^th^ to 90^th^ percentiles (whiskers) and were statistically evaluated by a linear mixed model grouped by severity (excluding low), with patient as random variable, corrected for age- and sex-dependency. CF: confounding factors; HI: hyperinflammatory; ICU: intensive care unit.

**Supplementary Figure 6: Patients in intensive care with COVID Hyperinflammation (HI) exhibit altered laboratory parameters and elevated IP-10.**

Four patients in our cohort were treated with intravenous methylprednisolone for COVID-Hyperinflammation; they are highlighted on the graph as HI 1-4. Laboratory parameters **a)** Ferritin (normal range 30-400μg/L), **b)** Procalcitonin (0.02-0.05μg/L), **c)** C-reactive protein (0-4mg/L) and **d)** D-Dimer (0-0.55mg/L) for HI patients versus COVID patients in intensive care unit who did not have features of HI (n’=11, black lines). **e)** Ferritin and **f)** plasma IP-10 concentration in HI (n’=4) and non-HI patients (n’=11, grey). Threshold (dotted grey line) demarcates e) upper limit of the normal range for ferritin as measured in our clinical laboratory (400μg/L) and f) directly below the lowest value detected for a COVID-HI patient. 5/11 non-HI COVID patients fell above ferritin threshold (labelled). **g)** Spearman correlation between plasma IP-10 and peak ferritin. n=95 n’=50. n=samples, n’=individuals. Shading in correlation plots represents 95% confidence intervals.

**Supplementary Figure 7: Flow cytometry gating strategies**.

**Data cleanup:** Pre-gating for all panels as indicated.

**Panel 1:** Gating strategy used to examine CD4^+^ T cells, CD8^+^ T cells, Tregs, B cells, NK cells and monocytes in human PBMCs by flow cytometry. In addition, naïve, effector and memory markers on T cells as well as NK cell subsets and CD25, CD103 and NKG2D expression were analysed. This gating strategy was used to examine relevant parameters in figures 1 and 5 and supplementary figures 2, 4 and 5.

**Panel 2:** Gating strategy used to examine naïve, effector and memory markers, CD25, CD103, CD38 and HLADR in CD4^+^ and CD8^+^ T cells and Tregs, Th1, Th17.1 and Th17 cells in CD4 T cells in human PBMCs by flow cytometry. This gating strategy was used to examine relevant parameters shown in figures 1,5 and 6, and supplementary figures 2,4 and 5.

**Panel 3:** Gating strategy used to examine Vδ1, Vδ2 and Vδx γδ T cells subsets and naïve and memory populations, PD1, CD103 and NKG2D expression within CD4^+^, CD8^+^ and γδ T cells in human PBMCs by flow cytometry. This gating strategy was used to examine relevant parameters shown in figures 1,5 and 6 and supplementary figures 2,4 and 5.

**Panel 4:** Gating strategy used to examine different B cell subpopulations: CD5^+^ B cells, plasmablasts, transitional B cells and naïve and memory B cells in human PBMCs by flow cytometry. This gating strategy was used to examine relevant parameters in figures 1 and 2 and supplementary figures 1 and 2.

**Panel 5:** Gating strategy used to examine proliferation status of Tregs, γδ T cells, naïve, effector, memory and EMRA CD4^+^ and CD8^+^ T cell subsets in human PBMCs by flow cytometry. This gating strategy was used to examine relevant parameters in figures 1 and 6 and supplementary figures 2,3,4 and 5.

**Panel 6:** Gating strategy used to examine T cells, B cells, NK cells, NKT cells, monocytes, neutrophils (and subtypes), basophils, and eosinophils in human whole blood by flow cytometry. Data from this panel was used to determine cell counts and is relevant to data shown in figures 1,2,3,4, 5 and 6 and supplementary figures 1,2,3,4 and 5.

**Panel 7:** Gating strategy used to examine PD1, LAG3, TIM3 and 2B4 expression by CD4^+^ and CD8^+^ T cells as well as by Th1, Th2, Th17 and Th17.1 and CD4 and CD8 memory populations in human whole blood by flow cytometry. This gating strategy was used to examine relevant parameters in figures 1,5 and 6 and supplementary figures 2,4 and 5.

**Panel 8:** Gating strategy used to examine eosinophils, neutrophils, basophils, monocytes (and subtypes), plasmacytoid DCs, myeloid DCs, CD1c^pos^ and CD1c^neg^ mDCs in human whole blood by flow cytometry. HLADR MFI and CD86 expression by monocyte and DC subsets were also analysed. This gating strategy was used to examine relevant parameters in figures 1,3 and 4 and supplementary figures 2 and 3.

**Panel 9:** Gating strategy used to examine cell cycle status of classical, intermediate and patrolling monocytes, CD14^low^ DCs and CD1c^pos^ and CD1c^neg^ mDCs. This gating strategy was used to examine relevant parameters in supplementary figure 3.

**Sorting gating strategy:** Gating strategy used to sort CD8 and CD4 T_EM_ for NanoString analysis.

